# Employment outcomes of people with Long Covid symptoms: community-based cohort study

**DOI:** 10.1101/2023.03.21.23287524

**Authors:** Daniel Ayoubkhani, Francesco Zaccardi, Koen B. Pouwels, A. Sarah Walker, Donald Houston, Nisreen A. Alwan, Josh Martin, Kamlesh Khunti, Vahé Nafilyan

## Abstract

**Background:** Evidence on the long-term employment consequences of SARS-CoV-2 infection is lacking. We used data from a large, community-based sample in the UK to estimate associations between Long Covid and subsequent employment outcomes.

**Methods:** This was an observational, longitudinal study using a pre-post design. We included survey participants from 3 February 2021 to 30 September 2022 when they were aged 16 to 64 years and not in full-time education. Using conditional logit modelling, we explored the time-varying relationship between Long Covid status ≥12 weeks after a first test-confirmed SARS-CoV-2 infection (reference: pre-infection) and labour market inactivity (neither working nor looking for work) or workplace absence lasting ≥4 weeks.

**Results:** Of 206,299 included participants (mean age 45 years, 54% female, 92% white), 15% were ever inactive in the labour market and 10% were ever long-term absent during follow-up. Compared with pre-infection, inactivity was higher in participants reporting Long Covid 30 to <40 weeks (adjusted odds ratio (aOR): 1.45; 95% CI: 1.17 to 1.81) or 40 to <52 weeks (1.34; 1.05 to 1.72) post-infection. Combining with official statistics on Long Covid prevalence, our estimates translate to 27,000 (95% CI: 6,000 to 47,000) working-age adults in the UK being inactive because of Long Covid in July 2022.

**Conclusions:** Long Covid is likely to have contributed to reduced levels of participation in the UK labour market, though it is unlikely to be the sole driver. Further research is required to quantify the contribution of other factors, such as indirect health effects of the pandemic.

## Introduction

A proportion of people infected with SARS[Z]CoV[Z]2 experience symptoms that persist for months or years after the acute infection, known as Long Covid. Common symptoms of Long Covid include fatigue, breathlessness, muscle and joint pain, cognitive impairment, and sleep disruption [1–3]. In January 2023, an estimated 2 million people in private households in the UK, or 3% of the population, reported long-term symptoms that they attributed to past SARS-CoV-2 infection; symptoms were having a detrimental impact on daily activities in 77% of these individuals and were most prevalent in the working-age population [4].

As the COVID-19 pandemic has progressed in the UK, the growing number of people infected with SARS-CoV-2 has coincided with rising levels of labour market inactivity, defined as neither being in work nor actively seeking work. By the end of 2022, the number of working-age people in the UK who were inactive had increased by over 350,000 since the start of the pandemic, with people aged 50 to 64 years contributing the majority of the increase; out of all possible reasons for inactivity, that due to long-term sickness grew the most quickly [5].

There is limited evidence on the relationship between Long Covid and subsequent changes in labour market participation. Given the widespread prevalence of Long Covid, this question has implications not only for the livelihood of individuals but also for the health of the macroeconomy, including income and earnings, labour market supply, productivity, tax receipts, benefit payments, and consumer demand. One study suggests that nearly half of UK firms had employees with Long Covid in 2022, and a quarter reported it as a substantial cause of long-term absence [6]. Long Covid may have resulted in long-term absence for 110,000 UK workers and the loss of 4.4 million working hours per week [7].

Despite the potential impact of Long Covid on global labour markets, there has been limited research based on individual-participant data. In one study, 45% of participants with Long Covid required a reduced work schedule and 22% were not working altogether due to their illness seven months post-infection [8]. In another analysis, 19% of participants with Long Covid were unable to work, 10% reported reduced working hours, and 37% said that their income had been affected [9]. Among participants with SARS-CoV-2, Long Covid symptoms have been found to be associated with 44% higher odds of not working and 27% lower odds of working full-time [10]. The largest study to date included 50,000 participants with SARS-CoV-2 in Germany, finding an average reduction in self-reported working capacity of 10.7% after a mean follow-up of 8.5 months post-infection [11]. While providing valuable insights, these studies are largely cross-sectional and descriptive in nature, lack robust control groups for comparison, and are based on convenience samples that may not be representative of the broader population. Meanwhile, inferential studies on Long Covid and employment outcomes are scarce, and limited sample sizes and follow-up time have precluded detailed analysis of population subgroups [7,12].

In this study, therefore, we used longitudinal data from a large, community-based probability sample to estimate associations between Long Covid and being out of employment and not looking for work, or experiencing long-term absence while in employment.

## Methods

### Study design and data

We performed an observational, longitudinal study using a pre-post design. The analysis included participants from the COVID-19 Infection Survey (CIS, ISRCTN21086382) [13], a longitudinal study of people aged ≥2 years from randomly sampled households (excluding hospitals, care homes, halls of residence, and prisons) across the UK. The study received ethical approval from the South Central Berkshire B Research Ethics Committee (20/SC/0195). Enrolment took place from 26 April 2020 to 31 January 2022, accruing over 530,000 participants (**Supplementary Table 1** reports enrolment rates). Following enrolment, >97% of participants provided written consent for monthly assessments for at least a year.

At each assessment, all participants provided a nose and throat self-swab for polymerase chain reaction (PCR) testing, while a sub-sample (initially approximately 10% of households but expanded from April 2021) also provided blood samples for antibody testing. Participants reported whether they had tested positive for SARS-CoV-2 or antibodies outside of the CIS, and whether they would describe themselves as currently experiencing Long Covid (defined as symptoms ≥4 weeks after a SARS-CoV-2 infection that could not be explained by another health condition). From April 2020 to June 2022, data collection was conducted via face-to-face interviews with study workers at participants’ homes. Study participants were transitioned to remote data collection from July 2022, whereby participants completed the survey questionnaire online or by telephone and returned samples through the post.

### Inclusion and exclusion criteria

We included monthly study assessments from 3 February 2021 (when data on Long Covid were first collected) to 30 September 2022 when participants responded to the Long Covid question, were aged 16 to 64 years, and were not in full-time education.

To ensure that a first positive swab for SARS-CoV-2 during the study period was likely to represent a first infection, we excluded participants with a first positive swab for SARS-CoV-2 (either a PCR test via study assessments or any swab test outside of the study, as self-reported by participants) at enrolment, as the timing of infection could not be determined for these participants. We also excluded participants with a positive spike-antibody blood test (excluding any tests after COVID-19 vaccination) or reported thinking they had COVID-19 ≥14 days before their first positive swab, as the first positive test may have represented a reinfection. To ensure we could fully observe participants’ self-reported Long Covid experience, we further excluded participants first testing positive before 11 November 2020 (12 weeks before the Long Covid survey question was implemented).

When analysing long-term absence, we excluded study assessments when participants were not in employment, as well as those before 1 October 2021 (when the UK Coronavirus Job Retention Scheme, also known as ‘furlough’, was in operation).

### Exposures

The time-varying exposure was past SARS-CoV-2 infection status (determined from CIS and self-reported positive swabs) by current Long Covid status (positive responses to the Long Covid survey question ≥12 weeks after a first positive swab): uninfected, infected in the past 12 weeks, infected ≥12 weeks ago without reporting Long Covid to date, infected ≥12 weeks ago and currently reporting Long Covid, and infected ≥12 weeks ago and previously reported Long Covid (potentially recovered or in remission). We therefore conceptualise the development of Long Covid symptoms as being the mediating factor between SARS-CoV-2 infection and labour market inactivity. Long Covid status was obtained from the following survey question: “Would you describe yourself as having ‘Long Covid’, that is, you are still experiencing symptoms more than 4 weeks after you first had COVID-19 that are not explained by something else?” Participants infected ≥12 weeks ago (with or without reporting Long Covid) were stratified by time since first positive test: 12 to <18, 18 to <24, 24 to <30, 30 to <40, 40 to <52, or ≥52 weeks.

### Outcomes

The outcomes of interest were labour market inactivity (excluding retirement, i.e., neither working nor looking for work, and not retired), and being absent from work for ≥4 weeks for any reason whilst in employment.

### Covariates

We considered as covariates a range of socio-demographic variables hypothesised to be related to both Long Covid and employment status: age at last birthday, sex, white or non-white ethnicity (sample sizes did not permit more detailed breakdowns), country/region of residence, area deprivation quintile group, and self-reported health/disability status (derived from the survey question: “Do you have any physical or mental health conditions or illnesses lasting or expected to last 12 months or more (excluding any long-lasting COVID-19 symptoms)?”). We also examined labour market variables: employment status, employment sector, Standard Occupational Classification (SOC) Major Group, and whether self-employed. All variables were measured at CIS enrolment and thus, by design, before SARS-CoV-2 infection for participants who were infected during follow-up.

### Statistical methods

We compared confounders between participants who ever or never reported Long Covid during follow-up using means and proportions for continuous and categorical variables, respectively. Absolute standardised differences >10% indicated imbalance between the groups [14].

We fitted conditional logit models to implicitly control for all measured and unmeasured time-invariant confounding, whereby each participant acts as their own control. To account for background labour market conditions over the study period, we adjusted for the calendar day of each study assessment (modelled as a restricted cubic spline with boundary knots at the 10^th^ and 90^th^ percentiles and a single internal knot at the median of the time distribution; sensitivity analyses included more knots), and interacted this with current age (restricted cubic spline, as for calendar day), sex, and self-reported health/disability status at CIS enrolment. We performed several sensitivity analyses for the primary outcome (labour market inactivity), detailed in **Supplementary Appendix 1**.

The conditional logit models were fitted using the ‘clogit’ function in R’s ‘survival’ package [15]. We reported results as adjusted odds ratios (aORs) and 95% confidence intervals (CIs), with pre-infection being the reference group for comparison. All statistical analyses were performed using R version 4.0.

### Heterogeneous effects

We tested for effect modification by socio-demographic characteristics (age, sex, ethnicity, underlying health/disability status, and area deprivation) at CIS enrolment. We also tested for heterogeneity by SARS-CoV-2 reinfection status at each follow-up assessment (using a pre-defined classification based on time since first positive swab and number of successive negative tests [16]), which may be associated with changes in Long Covid severity; and by mode of data collection to allow for differential exposure misclassification (CIS participants were 30% more likely to report Long Covid symptoms if responding remotely rather than face-to-face [17], perhaps due to stigma associated with the condition [18]). It was possible to test for effect modification by labour market attributes (employment sector, SOC Major Group, self-employment status) for long-term absence but not for economic inactivity, as these attributes can only be defined for people who are in work.

Modifiers were tested independently of one another. In each model, we included an interaction between calendar day of assessment and the modifier of interest to control for heterogeneity in background labour market conditions by the modifier. For each outcome and each modifier, statistically significant interactions were identified at the 5% level after performing Benjamini-Yekutieli [19] corrections to p-values to account for multiple comparisons across time-since-infection intervals and levels of modifiers.

### Population attributable risk

The underlying CIS data used in our study have also been used to produce official statistics on the population prevalence of Long Covid by inactivity status and time since infection [20]. By combining those population-level estimates with our aORs, we estimated the number of working-age adults in the UK who were inactive in July 2022 because of their Long Covid symptoms; that is, those reporting Long Covid who would have been working had they not been infected with SARS-CoV-2. A detailed description of these calculations can be found in **Supplementary Appendix 2**, but in summary:

1. From the published data, we calculated the odds of inactivity among people reporting Long Covid for each time-since-infection stratum
2. We divided the odds of inactivity by our aORs for people currently reporting Long Covid in each time-since-infection stratum to give an estimate of the counterfactual odds of inactivity (that is, the odds had those reporting Long Covid not been infected with SARS-CoV-2), assuming the statistical model is correct
3. We applied the counterfactual odds of inactivity to the total number of people reporting Long Covid in each time-since-infection stratum to give an estimate of the number of people reporting Long Covid who would have been inactive had they not been infected with SARS-CoV-2, assuming the statistical model is correct
4. For each time-since-infection stratum, we calculated the difference between the number of people reporting Long Covid who were inactive and the estimated number who would have been inactive had they not been infected with SARS-CoV-2; this gives an estimate of the inactivity attributable to Long Covid, assuming the statistical model is correct
5. We then summed the estimated attributable inactivity totals across time-since-infection strata
6. Finally, confidence intervals around the estimates were constructed using simulation, accounting for the uncertainty inherent in both inputs to our estimates: the number of people in the population reporting Long Covid by inactivity status; and the adjusted odds ratios for inactivity by time since first SARS-CoV-2 infection and current Long Covid status

## Results

### Characteristics of study participants

The analysed population comprised 206,299 participants aged 16 to 64 years who were not in full-time education, responded to the Long Covid question at least once between February 2021 and September 2022, and either had no evidence of SARS-CoV-2 by 31 September 2022 (52.6%) or had a first positive swab from 11 November 2020 (47.4%) **(Supplementary** Figure 1). Participants contributed a mean of 12.3 monthly assessments per participant.

147,895 participants were in employment during 1,155,207 study assessments from 1 October 2021 and were therefore included in the analysis of long-term absence. 97,751 participants tested positive for SARS-CoV-2 during follow-up and 8,440 (4.1% of the total, 8.6% of the infected) reported experiencing Long Covid ≥12 weeks post-infection.

Among participants testing positive for SARS-CoV-2, median follow-up from first positive swab to final assessment was 183 (interquartile range [IQR] 93 to 271) days, being longer for participants who reported Long Covid (313; 223 to 488) than those who did not (176; 85 to 258). Median time from first positive test was 35 (IQR: 12 to 59) days among assessments <12 weeks after infection; 184 (128 to 275) days for assessments ≥12 weeks after infection without reporting Long Covid to date; 207 (136 to 322) days for assessments when currently reporting Long Covid; and 310 (219 to 429) days for assessments after previously reported Long Covid.

Compared with participants infected with SARS-CoV-2 without ever reporting Long Covid during follow-up, those who reported Long Covid were on average older (46.3 versus 44.3 years) at CIS enrolment, and were more likely to be female (63.2% versus 55.1%), living in the most deprived area quintile group (14.9% versus 10.8%), living with a long-term health condition/disability (24.2% versus 16.2%), and not working and not looking for work (9.7% versus 6.6%) **(Table 1)**. Among infected participants who were employed at enrolment, those who reported Long Covid during follow-up were more likely to be working in teaching and education (16.0%, versus 12.8% of participants without reporting Long Covid) or national/local government (7.3% versus 6.3%), and employed in caring, leisure and other service occupations (10.3% versus 6.5%). A comparison of study participants who reported Long Covid during follow-up versus those not infected with SARS-CoV-2 can be found in **Supplementary Table 2**.

**Table 1.** Characteristics at enrolment of study participants ever infected with SARS-CoV-2 during follow-up, stratified by whether participants ever subsequently reported Long Covid

| Characteristic | Level | All infected participants<br>(*n*=97,751) | Never reported<br>Long Covid<br>(*n*=89,311) | Ever reported<br>Long Covid<br>(*n*=8,440) | Absolute<br>standardized<br>difference (%) |
| --- | --- | --- | --- | --- | --- |
| Age (years), mean (SD) |  | 44.5 (12.3) | 44.3 (12.3) | 46.3 (11.2) | 16.6 |
| Age group (*n*, %) | <16 years | 489 (0.5) | 474 (0.5) | 15 (0.2) | 20.8 |
|  | 16 to 24 years | 6,574 (6.7) | 6,197 (6.9) | 377 (4.5) |  |
|  | 25 to 34 years | 15,151 (15.5) | 14,176 (15.9) | 975 (11.6) |  |
|  | 35 to 49 years | 36,284 (37.1) | 32,984 (36.9) | 3,300 (39.1) |  |
|  | 50 to 64 years | 39,253 (40.2) | 35,480 (39.7) | 3,773 (44.7) |  |
| Sex (*n*, %) | Male | 43,201 (44.2) | 40,091 (44.9) | 3,110 (36.8) | 16.4 |
|  | Female | 54,550 (55.8) | 49,220 (55.1) | 5,330 (63.2) |  |
| Ethnic group (*n*, %) | White | 90,200 (92.3) | 82,326 (92.2) | 7,874 (93.3) | 4.3 |
|  | Non-white | 7,551 (7.7) | 6,985 (7.8) | 566 (6.7) |  |
| Country/region of residence (*n*, %) | North East England | 3,549 (3.6) | 3,160 (3.5) | 389 (4.6) | 13.6 |
|  | North West England | 11,135 (11.4) | 10,073 (11.3) | 1,062 (12.6) |  |
|  | Yorkshire and the Humber | 8,088 (8.3) | 7,349 (8.2) | 739 (8.8) |  |
|  | East Midlands | 6,142 (6.3) | 5,546 (6.2) | 596 (7.1) |  |
|  | West Midlands | 7,061 (7.2) | 6,388 (7.2) | 673 (8.0) |  |
|  | East of England | 8,723 (8.9) | 7,921 (8.9) | 802 (9.5) |  |
|  | London | 17,253 (17.6) | 16,029 (17.9) | 1,224 (14.5) |  |
|  | South East England | 11,853 (12.1) | 10,922 (12.2) | 931 (11.0) |  |
|  | South West England | 7,482 (7.7) | 6,876 (7.7) | 606 (7.2) |  |
|  | Scotland | 8,268 (8.5) | 7,590 (8.5) | 678 (8.0) |  |
|  | Wales | 4,897 (5.0) | 4,442 (5.0) | 455 (5.4) |  |
|  | Northern Ireland | 3,300 (3.4) | 3,015 (3.4) | 285 (3.4) |  |
| Area deprivation quintile group (*n*, %) | 1 (most deprived) | 10,889 (11.1) | 9,629 (10.8) | 1,260 (14.9) | 14.7 |
|  | 2 | 16,510 (16.9) | 14,889 (16.7) | 1,621 (19.2) |  |
|  | 3 | 20,408 (20.9) | 18,645 (20.9) | 1,763 (20.9) |  |
|  | 4 | 23,441 (24.0) | 21,608 (24.2) | 1,833 (21.7) |  |
|  | 5 (least deprived) | 26,503 (27.1) | 24,540 (27.5) | 1,963 (23.3) |  |
| Self-reported health/disability status (*n*, %) | No long-term health conditions | 81,211 (83.1) | 74,813 (83.8) | 6,398 (75.8) | 20.4 |
|  | Health conditions without impact to day-to-day activities | 9,358 (9.6) | 8,422 (9.4) | 936 (11.1) |  |
|  | Day-to-day activities limited a little by health conditions | 4,572 (4.7) | 3,906 (4.4) | 666 (7.9) |  |
|  | Day-to-day activities limited a lot by health conditions | 2,610 (2.7) | 2,170 (2.4) | 440 (5.2) |  |

**Table 1 (continued)**
| Characteristic | Level | All infected participants<br>(n=97,751) | Never reported Long Covid<br>(n=89,311) | Ever reported Long Covid<br>(n=8,440) | Absolute standardized difference (%) |
| --- | --- | --- | --- | --- | --- |
| Employment status (n, %) | Employed | 79,121 (80.9) | 72,356 (81.0) | 6,765 (80.2) | 16.8 |
|  | Unemployed | 1,885 (1.9) | 1,706 (1.9) | 179 (2.1) |  |
|  | Not working and not looking for work | 6,759 (6.9) | 5,939 (6.6) | 820 (9.7) |  |
|  | Retired | 6,747 (6.9) | 6,209 (7.0) | 538 (6.4) |  |
|  | Student | 3,239 (3.3) | 3,101 (3.5) | 138 (1.6) |  |
| Employment sector, among participants in employment (n, %) | Teaching and education | 10,374 (13.1) | 9,289 (12.8) | 1,085 (16.0) | 14.1 |
|  | Health care | 7,315 (9.2) | 6,676 (9.2) | 639 (9.4) |  |
|  | Social care | 1,988 (2.5) | 1,782 (2.5) | 206 (3.0) |  |
|  | Transport | 2,527 (3.2) | 2,305 (3.2) | 222 (3.3) |  |
|  | Retail and wholesale | 4,781 (6.0) | 4,370 (6.0) | 411 (6.1) |  |
|  | Hospitality | 2,040 (2.6) | 1,846 (2.6) | 194 (2.9) |  |
|  | Food production, agriculture and farming | 1,150 (1.5) | 1,053 (1.5) | 97 (1.4) |  |
|  | Personal services | 856 (1.1) | 786 (1.1) | 70 (1.0) |  |
|  | Information technology and communication | 4,906 (6.2) | 4,604 (6.4) | 302 (4.5) |  |
|  | Financial services | 5,795 (7.3) | 5,430 (7.5) | 365 (5.4) |  |
|  | Manufacturing and construction | 6,793 (8.6) | 6,228 (8.6) | 565 (8.4) |  |
|  | Civil service and local government | 5,031 (6.4) | 4,540 (6.3) | 491 (7.3) |  |
|  | Armed forces | 250 (0.3) | 232 (0.3) | 18 (0.3) |  |
|  | Arts, entertainment and recreation | 1,599 (2.0) | 1,479 (2.0) | 120 (1.8) |  |
|  | Other | 10,305 (13.0) | 9,487 (13.1) | 818 (12.1) |  |
|  | Unknown | 13,411 (16.9) | 12,249 (16.9) | 1,162 (17.2) |  |
| SOC Major Group, among participants in employment (n, %) | Managers, directors and senior officials | 6,990 (8.8) | 6,456 (8.9) | 534 (7.9) | 18.4 |
|  | Professional occupations | 18,521 (23.4) | 17,143 (23.7) | 1,378 (20.4) |  |
|  | Associate professional and technical occupations | 12,488 (15.8) | 11,509 (15.9) | 979 (14.5) |  |
|  | Administrative and secretarial occupations | 8,903 (11.3) | 8,093 (11.2) | 810 (12.0) |  |
|  | Skilled trades occupations | 4,894 (6.2) | 4,457 (6.2) | 437 (6.5) |  |
|  | Caring, leisure and other service occupations | 5,421 (6.9) | 4,727 (6.5) | 694 (10.3) |  |
|  | Sales and customer service occupations | 3,380 (4.3) | 3,065 (4.2) | 315 (4.7) |  |
|  | Process, plant and machine operatives | 2,380 (3.0) | 2,166 (3.0) | 214 (3.2) |  |
|  | Elementary occupations | 3,253 (4.1) | 2,927 (4.0) | 326 (4.8) |  |
|  | Unknown | 12,891 (16.3) | 11,813 (16.3) | 1,078 (15.9) |  |
| Self-employment status, among participants in employment (n, %) | Employee | 72,616 (91.8) | 66,412 (91.8) | 6,204 (91.7) | 0.3 |
|  | Self-employed | 6,505 (8.2) | 5,944 (8.2) | 561 (8.3) |  |

Notes: SD: standard deviation; SOC: Standard Occupational Classification. Area deprivation was based on the English Indices of Deprivation 2019, the Welsh Index of Multiple Deprivation 2019, the Scottish Index of Multiple Deprivation 2020, and the Northern Ireland Multiple Deprivation Measure 2017. Health conditions were self-reported rather than clinically diagnosed based on the survey question: "Do you have any physical or mental health conditions or illnesses lasting or expected to last 12 months or more (excluding any long-lasting COVID-19 symptoms)?"

### Labour market inactivity

31,248 study participants (15.1%) were ever inactive (excluding retired) during follow-up. Irrespective of timing, 17.7% of participants who ever reported Long Covid during follow-up were ever inactive, compared with 13.4% of those infected with SARS-CoV-2 during follow-up without reporting Long Covid. Participants were inactive for 10.3% of study assessments while currently reporting Long Covid, compared with 5.8% of assessments ≥12 weeks post-infection without reporting Long Covid and 7.7% of assessments after previously reporting Long Covid **(Table 2)**.

**Table 2.** Percentage of monthly study assessments spent in each employment status, stratified by exposure status.

| Exposure status | Total number of assessments in exposure state | Percentage of assessments in each employment state |  |  |  |
| --- | --- | --- | --- | --- | --- |
|  |  | Employed | Unemployed | Inactive (exc. retired) | Retired |
| Uninfected | 1,972,041 | 77.5 | 2.0 | 8.2 | 12.2 |
| Infected <12 weeks ago | 212,976 | 83.7 | 1.3 | 6.3 | 8.7 |
| Infected ≥12 weeks ago, never reported Long Covid to date | 310,517 | 85.4 | 1.3 | 5.8 | 7.4 |
| Infected ≥12 weeks ago, currently reporting Long Covid | 25,043 | 80.8 | 1.5 | 10.3 | 7.4 |
| Infected ≥12 weeks ago, previously reported Long Covid | 27,041 | 83.6 | 1.2 | 7.7 | 7.4 |

Compared with the pre-infection period, inactivity was less common in the first 12 weeks post-infection (aOR: 0.95; 95% CI: 0.91 to 0.99) and 12 to <18 weeks post-infection (without reporting Long Covid: 0.87 [0.82 to 0.93]; while reporting Long Covid: 0.83 [0.68 to 1.00]; previously reported Long Covid: 0.60 [0.36 to 1.00]) **(Figure 1)**. Beyond 18 weeks post-infection, there was no evidence of differences in the odds of inactivity compared with pre-infection for participants who had not reported Long Covid to date or had previously reported Long Covid. Conversely, participants currently reporting Long Covid 30 to <40 or 40 to <52 weeks post-infection were significantly more likely to be inactive compared with pre-infection, with aORs of 1.45 (1.17 to 1.81) and 1.34 (1.05 to 1.72), respectively.

**Figure 1.**
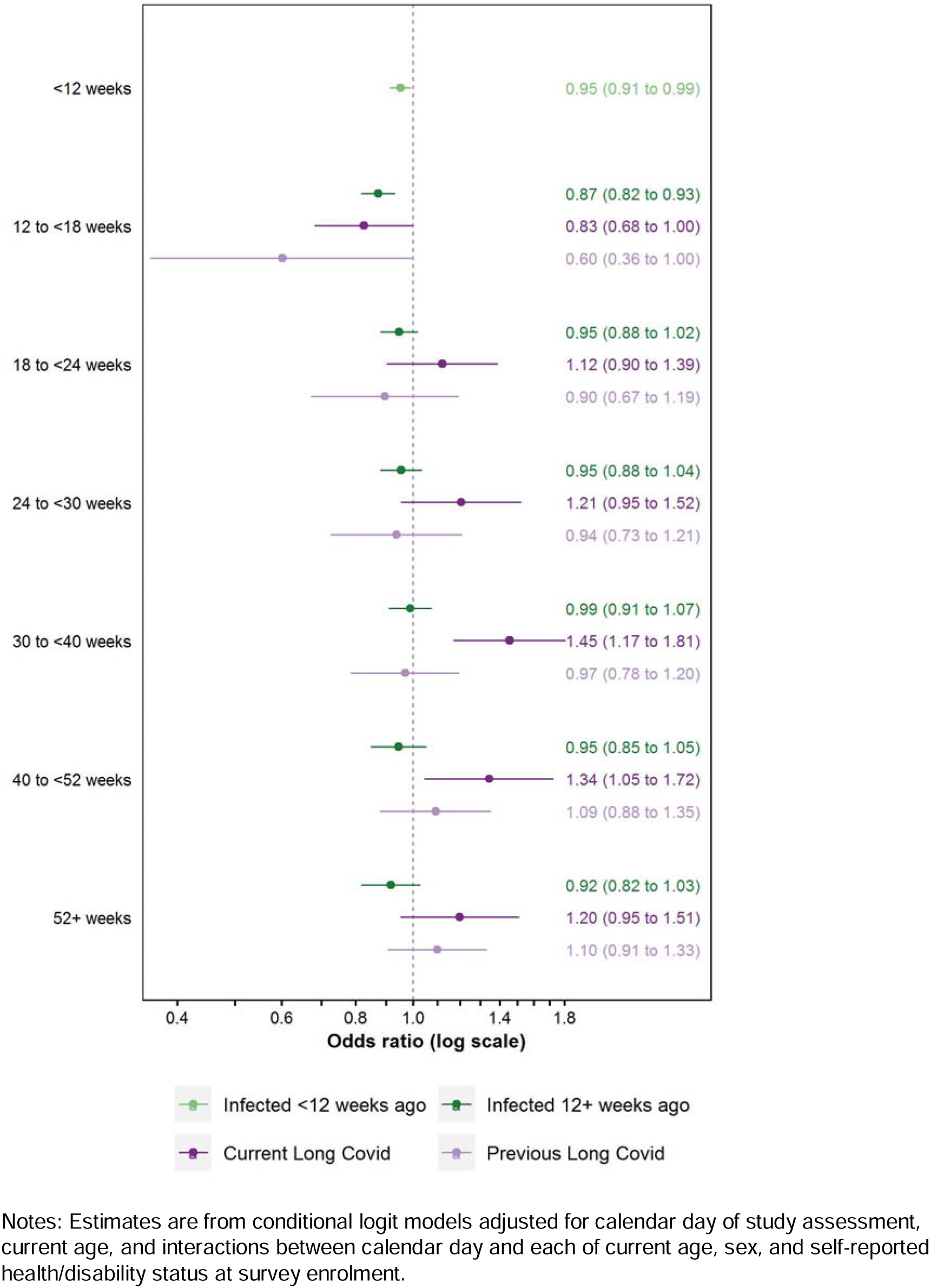
Adjusted odds ratios for inactivity (excluding retirement) compared with the pre-infection period, by time since SARS-CoV-2 infection and Long Covid status

Applying these aORs to published official statistics on the population prevalence of Long Covid by inactivity status and time since infection [20], an estimated 27,000 (95% CI: 6,000 to 47,000) working-age non-students were inactive (excluding retirement) because of their Long Covid symptoms in July 2022. Of these people, 16,000 (7,000 to 24,000) were aged 50 to 64 years.

There was no evidence of heterogeneity in the relationship between Long Covid and inactivity by socio-demographic characteristics, SARS-CoV-2 reinfection status, or data collection mode in any time-since-infection interval **(Supplementary Table 3)**. Despite not reaching the 5% threshold for statistical significance, aORs were consistently numerically higher for participants reporting Long Covid aged 50 to 64 years than for those aged 16 to 49 years **(Figure 2)**; the former group had elevated odds of inactivity for all time intervals from 24 weeks post-infection compared with pre-infection, peaking at 30 to <40 weeks (aOR: 1.71; 95% CI: 1.28 to 2.29).

**Figure 2.**
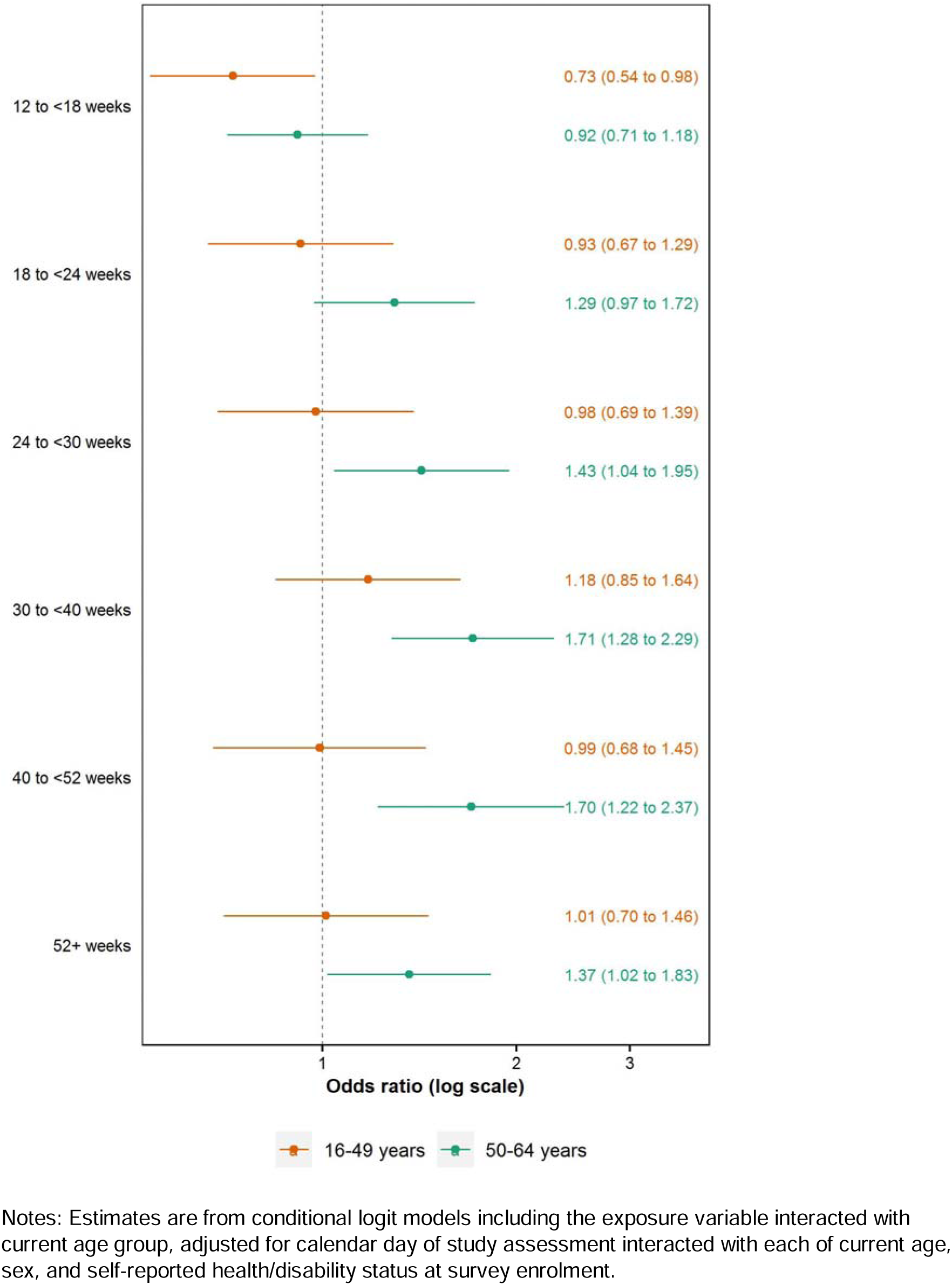
Adjusted odds ratios for inactivity (excluding retirement) for participants currently reporting Long Covid compared with the pre-infection period, by time since SARS-CoV-2 infection and age group

In sensitivity analysis, similar results were obtained when restricting the population to participants testing positive for SARS-CoV-2, excluding assessments when participants were retired, and increasing the number of internal knots in the splines for calendar time and age **(Supplementary Appendix 1)**.

### Long-term absence

Of 147,895 participants in employment from 1 October 2021, 14,493 (9.8%) reported long-term (≥4 weeks) absence during follow-up. Irrespective of timing, long-term absence was experienced by 13.1% of participants who reported Long Covid during follow-up, compared with 9.8% of those infected with SARS-CoV-2 without reporting Long Covid. Participants were long-term absent for 4.5% of study assessments while currently reporting Long Covid, compared with 3.1% of assessments before SARS-CoV-2 infection, 3.1% within the first 12 weeks post-infection, 3.2% ≥12 weeks post-infection without reporting Long Covid to date, and 2.8% after previously reporting Long Covid.

Compared with the pre-infection period, SARS-CoV-2 infection <12 weeks previously was associated with an increased likelihood of long-term absence (aOR: 1.09; 95% CI: 1.02 to 1.17); as too was reporting Long Covid 18 to <24 or 24 to <30 weeks post-infection, with aORs of 1.40 (1.04 to 1.90) and 1.45 (1.03 to 2.04), respectively **(Figure 3)**. Conversely, infection in the past 12 to <18 weeks without reporting Long Covid to date (0.84; 0.76 to 0.93), or being 40 to <52 weeks (0.70; 0.49 to 1.00) or ≥52 weeks (0.59; 0.40 to 0.86) after infection having previously reported Long Covid, were both associated with reduced odds of long-term absence relative to pre-infection.

**Figure 3.**
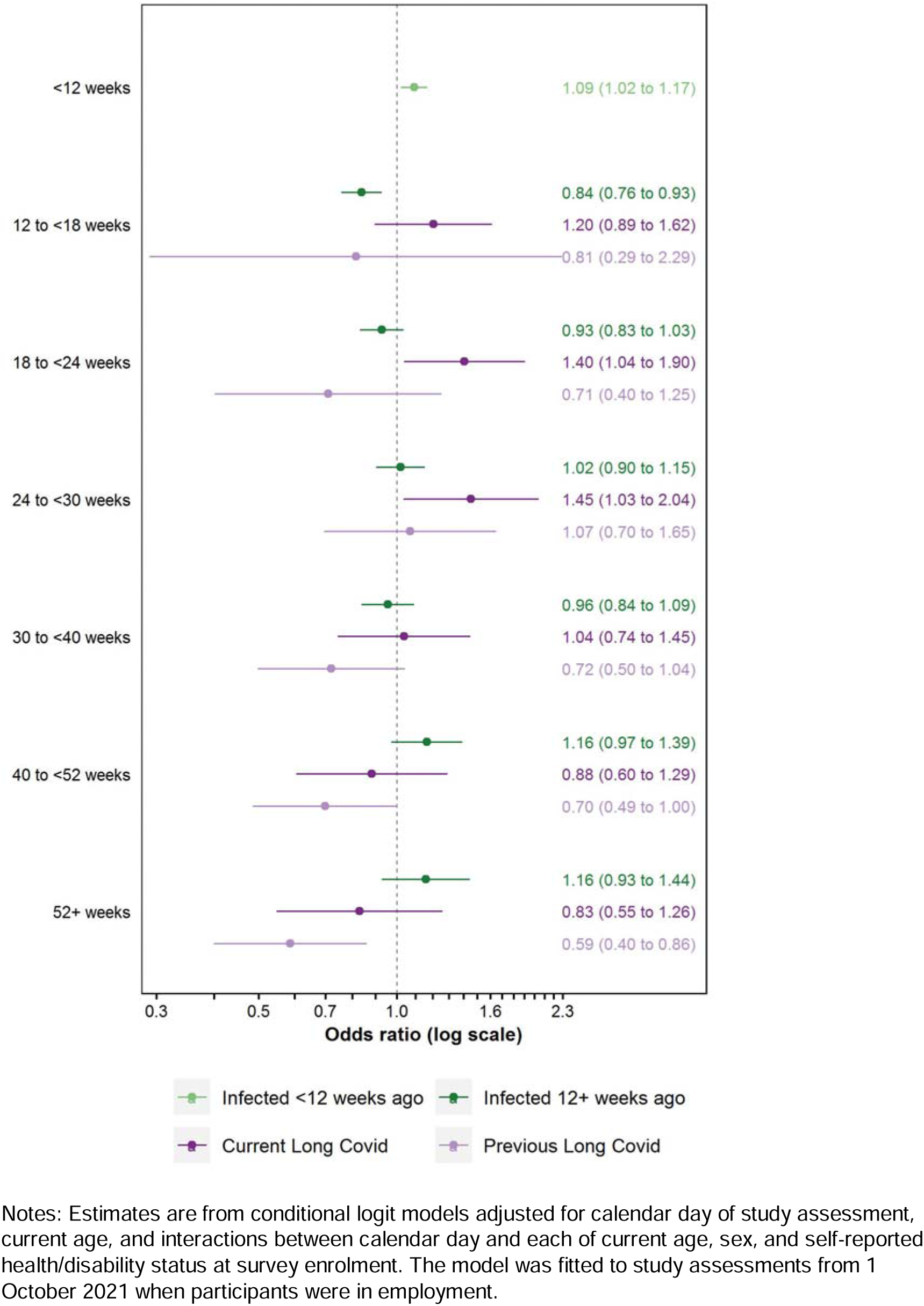
Adjusted odds ratios for long-term (≥4 weeks) absence compared with the pre-infection period, by time since SARS-CoV-2 infection and Long Covid status

There was no evidence of heterogeneity in the relationship between Long Covid and long-term absence for any effect modifier in any time-since-infection interval **(Supplementary Table 4)**, except for the presence of underlying health conditions 18 to <24 weeks post-infection (p=0.02). Participants without health conditions who reported Long Covid in this interval were more likely to be long-term absent compared with pre-infection (aOR: 1.96; 95% CI: 1.38 to 2.78), while there was no evidence of difference between the pre– and post-infection periods for participants with health conditions (0.58; 0.31 to 1.10).

## Discussion

### Principal findings

Among working-age individuals, reporting Long Covid after SARS-CoV-2 infection was associated with increased odds of being out of employment and not looking for work compared with the period before infection. This translates to an estimated 27,000 people not participating in the labour market due to their Long Covid symptoms.

The period of greatest excess risk of labour market inactivity (excluding retirement) among people reporting Long Covid was 30 to <40 weeks post-infection, with 45% higher odds relative to pre-infection. Compared with pre-infection, individuals reporting Long Covid were also at increased risk of long-term absence 18 to <30 weeks after infection, but not beyond 30 weeks. There was no evidence of such temporal relationships for people infected with SARS-CoV-2 without reporting Long Covid. Thus, is appears that SARS-CoV-2 infection is associated with downstream labour market implications, and the causal pathway by which this occurs is via long-term symptomology.

There was no evidence of heterogeneity in the relationship between Long Covid and inactivity or long-term absence by any of the explored effect modifiers, other than the presence of underlying health conditions 18 to <24 weeks post-infection when considering long-term absence. However, we cannot exclude this single statistically significant result being a chance finding.

### Findings in context

Our results complement and add to those from a limited number of longitudinal studies examining the employment outcomes of COVID-19, all with smaller sample sizes and shorter follow-up, from which the findings to date are mixed. Among 17,000 UK participants recruited via social and traditional media, SARS-CoV-2 was associated with a five-fold increase in the odds of sickness absence beyond the acute phase of the infection, but there was no evidence of a relationship between COVID-19 and inadequate household income in the long-term [12]. Among 36,000 UK Household Longitudinal Study (UKHLS) respondents, Long Covid was associated with reduced working hours but not being out of employment, with the former appearing to dissipate after six months [7].

Our principal finding of an association between Long Covid and inactivity is coherent with the broader labour market landscape in the UK, which has been characterised by rising levels of inactivity throughout the pandemic, primarily driven by people aged 50 to 64 years and long-term sickness [21]. However, it seems unlikely that Long Covid is the sole, or even main, driver of this trend towards inactivity. The number of working-age adults who were inactive due to ill-health had been gradually rising since early 2019, nearly a year before the emergence of COVID-19 [22]. Furthermore, persistently increasing levels of inactivity during the pandemic has not been commonplace internationally, despite Long Covid having a global burden [2]. The UK is among only nine of 38 Organisation for Economic Co-operation and Development (OECD) member states for which the inactivity rate among people aged 15 to 64 years was higher in the third quarter of 2022 than three years earlier, and one of only six for which the rate continued to rise over the latest four quarters [23].

To contextualise our estimated attributable risk of 27,000 people inactive because of their Long Covid symptoms, this represents just 0.5% of total inactivity (excluding retirement) among working-age non-students in the UK in July 2022 [21], and 13% of those reporting Long Covid [20]. This suggests that the majority of inactive people with Long Covid may have been absent from the labour market even if they had not been infected with SARS-CoV-2, for example due to other health conditions. Other studies suggest that 80,000 [24] to 96,000 [25] people might have left employment directly because of Long Covid by March 2022. These numbers are higher than our estimate as they represent cumulative exits from employment rather than point-in-time measures of inactivity.

We observed an inverted U-shaped relationship between Long Covid and inactivity (excluding retirement) over time, with the odds ratios peaking at 30 to <40 weeks post-infection before subsiding thereafter. This observation, coupled with lack of evidence of a relationship between time since infection and inactivity for individuals who previously had Long Covid symptoms, suggests that some people who left employment while experiencing Long Covid may have later returned to work (either in remission or with residual symptoms). Indeed, further analysis of the study data revealed that of the 8,440 participants who reported Long Covid during follow-up, 4.6% transitioned into inactivity whilst reporting persistent symptoms, but 27.3% of these had returned to employment by the end of the study period. This may have been facilitated by there being 1.2 million vacancies in the UK during the fourth quarter of 2022, 40% more than before the pandemic [26]. Furthermore, despite people with Long Covid in the UK potentially being eligible to claim unemployment– and disability-related social security benefits (universal credit, employment and support allowance, and personal independence payments), some may have been encouraged back to work due to financial pressures and the increased cost of living; the rate of consumer price inflation was high in the UK during 2022, peaking at 11.1% in October 2022 compared with 12 months earlier [27].

Irrespective of Long Covid status, individuals experienced reduced odds of inactivity in the first 18 weeks following SARS-CoV-2 infection, compared with pre-infection. Given that the risk of long-term absence was elevated in the first 12 weeks post-infection, possibly reflecting the effects of acute COVID-19, the short-term reduction in the odds of inactivity after this period may reflect a reduced propensity to leave employment during, or shortly after, a period of acute illness. We also cannot rule out some degree of reverse causality (that is, reduced exposure to SARS-CoV-2 infection, and thus Long Covid, after leaving employment).

In the longer-term, our finding that the risk of absence levels off after 30 weeks post-infection may partly reflect people returning to work (including those with persistent illness) upon completion of their 28-week period of statutory sick pay. Continuing to work while sick, so called “presenteeism”, has been linked to reduced productivity [28] and increased rates of future sickness absence [29]. However, we also cannot rule out a survivorship effect: study participants remaining at risk of long-term absence beyond 30 weeks post-infection were those who had not yet left the workforce, including due to ill-health.

### Strengths and limitations

To the authors’ knowledge, this is the largest longitudinal study to date to examine the relationship between Long Covid and employment outcomes, with over 200,000 participants included in the analysis. Selection bias was minimised by selecting households at random from national address lists, while prospective data collection meant that responses were not affected by recall effects (such as overestimating the duration of symptoms or approximating the timing of changes in employment status). The conditional logit modelling approach controlled for all observed and unobserved time-invariant confounders.

Our study also has limitations. We cannot rule out residual confounding by unmeasured time-varying factors, and reverse causality may have affected the estimated relationships between Long Covid and labour market status to some extent. Bias may also be present if loss to follow-up after SARS-CoV-2 infection is related to employment changes. This would partly explain our finding of an inverse U-shaped relationship between Long Covid and inactivity if participants who were out of work for long durations (perhaps due to ill-health) were also most likely to leave the study.

Absolute outcome rates should be interpreted with caution because people who enrol onto the CIS may not be representative of the broader working-age population in terms of labour market attributes, including unobserved traits such as motivation and aptitudes. For example, CIS participants are less likely to be unemployed but more likely to be inactive than the general population **(Supplementary** Figure 2). However, this is unlikely to have biased the estimated relative (pre-versus post-infection) risks that are the main focus of the analysis.

We did not have data on working hours, so we were unable to investigate the relationship between Long Covid and reduced working time. It was also not possible to distinguish long-term absence due to sickness from that due to other reasons, for example maternity/paternity leave. However, the latter are unlikely to be influenced by past SARS-CoV-2 infection, so we expect the impact on the estimated relative risks to be small.

Total inactivity attributable to Long Covid may be underestimated because it does not include the effects of early retirement, transitioning to unemployment (not working but looking for work and available to start), or students choosing to remain in education instead of finding work. However, we expect these effects to be small because: there were 27,000 fewer working-age retirees in the UK in the final quarter of 2022 compared with 2019, before the onset of the pandemic [21]; the unemployment rate in the UK dropped to 3.5% during 2022, a historic low since comparable records began in 1992 [21]; and the population prevalence of Long Covid is considerably lower among people aged <25 years (a group comprising the majority of students) than in older people [4]. Furthermore, our estimates do not consider any indirect impacts on employment, such as family members of people with Long Covid reducing their working hours or leaving employment to take on caring responsibilities; the presence and extent of such effects remain unknown.

Long Covid status was self-reported by study participants so exposure misclassification is possible (for example, some participants’ symptoms may have been caused by a medical condition unrelated to COVID-19). However, there is currently no biological test for Long Covid, and it remains a “diagnosis of exclusion”. Case ascertainment based on recorded clinical diagnoses is therefore likely to lack sensitivity due to under-presentation, under-diagnosing, and under-coding [30].

Finally, complete confounding between calendar time and SARS-CoV-2 variant of infection meant that we could not assess whether the relationship between Long Covid and inactivity differed by variant.

### Conclusions

Long Covid is associated with labour market inactivity among working-age people, with the greatest excess risk occurring 30 to <40 weeks after SARS-CoV-2 infection compared with pre-infection. Long Covid is also associated with long-term absence 18 to <30 weeks after infection. It is therefore likely that Long Covid has contributed to reduced participation in the UK labour market. At the same time, it is unlikely to have been the sole driver of inactivity, with fewer than 30,000 working-age adults estimated to be inactive because of their Long Covid symptoms in July 2022.

The relative contribution of factors besides Long Covid to reduced levels of labour market participation (such as indirect health effects of the pandemic and extended healthcare waiting lists) remains unknown, and further research is required. Future studies with longer follow-up could also provide insights on a broader range of socio-economic outcomes following SARS-CoV-2 infection, such as income and earnings, social security benefit claims, and socio-economic position.

## Acknowledgements

DA thanks Tarnjit Khera at the Office for National Statistics (ONS) for peer review of the analytical code. DA, KK and FZ are supported by the National Institute for Health Research (NIHR) Applied Research Collaboration East Midlands (ARC EM). KK and FZ are supported by the NIHR Leicester Biomedical Research Centre (BRC). KBP and ASW are supported by the NIHR Health Protection Research Unit in Healthcare Associated Infections and Antimicrobial Resistance (NIHR200915), a partnership between the UK Health Security Agency (UKHSA) and the University of Oxford. KBP is supported by the Huo Family Foundation. ASW is supported by the NIHR Oxford Biomedical Research Centre and is an NIHR Senior Investigator. The views expressed are those of the authors and are not necessarily those of the National Health Service, NIHR, the Department of Health and Social Care, UKHSA, or the Bank of England or any of its committees. For the purpose of open access, the authors have applied a Creative Commons Attribution (CC BY) licence to any Author Accepted Manuscript version arising.

## Footnotes

### Ethics

The CIS received ethical approval from the South Central Berkshire B Research Ethics Committee (20/SC/0195).

### Patient and public involvement

NAA has lived experience of Long Covid.

### Data availability

De-identified study data are available to accredited researchers in the ONS Secure Research Service (SRS) under part 5, chapter 5 of the Digital Economy Act 2017. For further information about accreditation, contact or visit: https://www.ons.gov.uk/aboutus/whatwedo/statistics/requestingstatistics/approvedresearcher scheme

## Funding

The CIS is funded by the Department of Health and Social Care with in-kind support from the Welsh Government, the Department of Health on behalf of the Northern Ireland Government, and the Scottish Government. There was no dedicated funding for this analysis of CIS data.

### Author contributions

DA and VN conceptualised and designed the study. DA prepared the study data and performed the statistical analysis. All authors contributed to interpretation of the results. DA was responsible for the first draft of the manuscript. All authors contributed to critical revision of the manuscript. All authors approved the final manuscript.

### Competing interests

All authors have completed the ICMJE uniform disclosure form at http://www.icmje.org/disclosure-of-interest/ and declare: no support from any organisation for the submitted work; and no financial relationships with any organisations that might have an interest in the submitted work in the previous three years. KK chairs the Long Covid research-funded group reporting to the Chief Medical Officer, chairs the Ethnicity Subgroup of the UK Scientific Advisory Group for Emergencies (SAGE), and is a Member of SAGE. KK and NAA are co-investigators on the NIHR-funded STIMULATE-ICP study. NAA is a co-investigator on the NIHR-funded HI-COVE study.

**Supplementary Table 1.** Characteristics at enrolment of study participants who were either never infected with SARS-CoV-2 during follow-up, or who were infected and subsequently ever reported Long Covid

| Characteristic | Level | All participants<br>(*n*=116,988) | Never infected<br>(*n*=108,548) | Ever reported<br>Long Covid<br>(*n*=8,440) | Absolute<br>standardized<br>difference (%) |
| --- | --- | --- | --- | --- | --- |
| Age (years), mean (SD) |  | 45.2 (13.7) | 45.1 (13.9) | 46.3 (11.2) | 8.9 |
| Age group (*n*, %) | <16 years | 649 (0.6) | 634 (0.6) | 15 (0.2) | 37.9 |
|  | 16 to 24 years | 10,484 (9.0) | 10,107 (9.3) | 377 (4.5) |  |
|  | 25 to 34 years | 18,711 (16.0) | 17,736 (16.3) | 975 (11.6) |  |
|  | 35 to 49 years | 33,509 (28.6) | 30,209 (27.8) | 3,300 (39.1) |  |
|  | 50 to 64 years | 53,635 (45.8) | 49,862 (45.9) | 3,773 (44.7) |  |
| Sex (*n*, %) | Male | 54,769 (46.8) | 51,659 (47.6) | 3,110 (36.8) | 21.9 |
|  | Female | 62,219 (53.2) | 56,889 (52.4) | 5,330 (63.2) |  |
| Ethnic group (*n*, %) | White | 106,958 (91.4) | 99,084 (91.3) | 7,874 (93.3) | 7.5 |
|  | Non-white | 10,030 (8.6) | 9,464 (8.7) | 566 (6.7) |  |
| Country/region of residence (*n*, %) | North East England | 3,943 (3.4) | 3,554 (3.3) | 389 (4.6) | 17.7 |
|  | North West England | 12,127 (10.4) | 11,065 (10.2) | 1,062 (12.6) |  |
|  | Yorkshire and the Humber | 8,789 (7.5) | 8,050 (7.4) | 739 (8.8) |  |
|  | East Midlands | 7,495 (6.4) | 6,899 (6.4) | 596 (7.1) |  |
|  | West Midlands | 8,553 (7.3) | 7,880 (7.3) | 673 (8.0) |  |
|  | East of England | 10,847 (9.3) | 10,045 (9.3) | 802 (9.5) |  |
|  | London | 21,420 (18.3) | 20,196 (18.6) | 1,224 (14.5) |  |
|  | South East England | 14,553 (12.4) | 13,622 (12.5) | 931 (11.0) |  |
|  | South West England | 9,264 (7.9) | 8,658 (8.0) | 606 (7.2) |  |
|  | Scotland | 10,266 (8.8) | 9,588 (8.8) | 678 (8.0) |  |
|  | Wales | 6,427 (5.5) | 5,972 (5.5) | 455 (5.4) |  |
|  | Northern Ireland | 3,304 (2.8) | 3,019 (2.8) | 285 (3.4) |  |
| Area deprivation quintile group (*n*, %) | 1 (most deprived) | 14,449 (12.4) | 13,189 (12.2) | 1,260 (14.9) | 7.2 |
|  | 2 | 21,078 (18.0) | 19,457 (17.9) | 1,621 (19.2) |  |
|  | 3 | 25,228 (21.6) | 23,465 (21.6) | 1,763 (20.9) |  |
|  | 4 | 27,163 (23.2) | 25,330 (23.3) | 1,833 (21.7) |  |
|  | 5 (least deprived) | 29,070 (24.8) | 27,107 (25.0) | 1,963 (23.3) |  |
| Self-reported health/disability status (*n*, %) | No long-term health conditions | 92,758 (79.3) | 86,360 (79.6) | 6,398 (75.8) | 8.6 |
|  | Health conditions without impact to day-to-day activities | 11,539 (9.9) | 10,603 (9.8) | 936 (11.1) |  |
|  | Day-to-day activities limited a little by health conditions | 7,131 (6.1) | 6,465 (6.0) | 666 (7.9) |  |
|  | Day-to-day activities limited a lot by health conditions | 5,560 (4.8) | 5,120 (4.7) | 440 (5.2) |  |

**Supplementary Table 1 (continued)**
| Characteristic | Level | All participants<br>(n=116,988) | Never infected<br>(n=108,548) | Ever reported<br>Long Covid<br>(n=8,440) | Absolute<br>standardized<br>difference (%) |
| --- | --- | --- | --- | --- | --- |
| Employment status (n, %) | Employed | 85,271 (72.9) | 78,506 (72.3) | 6,765 (80.2) | 23.9 |
|  | Unemployed | 3,325 (2.8) | 3,146 (2.9) | 179 (2.1) |  |
|  | Not working and not looking for work | 10,637 (9.1) | 9,817 (9.0) | 820 (9.7) |  |
|  | Retired | 12,984 (11.1) | 12,446 (11.5) | 538 (6.4) |  |
|  | Student | 4,771 (4.1) | 4,633 (4.3) | 138 (1.6) |  |
| Employment sector, among<br>participants in employment (n, %) | Teaching and education | 8,876 (10.4) | 7,791 (9.9) | 1,085 (16.0) | 18.0 |
|  | Health care | 7,348 (8.6) | 6,709 (8.5) | 639 (9.4) |  |
|  | Social care | 2,152 (2.5) | 1,946 (2.5) | 206 (3.0) |  |
|  | Transport | 2,916 (3.4) | 2,694 (3.4) | 222 (3.3) |  |
|  | Retail and wholesale | 5,826 (6.8) | 5,415 (6.9) | 411 (6.1) |  |
|  | Hospitality | 2,606 (3.1) | 2,412 (3.1) | 194 (2.9) |  |
|  | Food production, agriculture and farming | 1,428 (1.7) | 1,331 (1.7) | 97 (1.4) |  |
|  | Personal services | 1,043 (1.2) | 973 (1.2) | 70 (1.0) |  |
|  | Information technology and communication | 5,808 (6.8) | 5,506 (7.0) | 302 (4.5) |  |
|  | Financial services | 6,156 (7.2) | 5,791 (7.4) | 365 (5.4) |  |
|  | Manufacturing and construction | 7,717 (9.0) | 7,152 (9.1) | 565 (8.4) |  |
|  | Civil service and local government | 5,403 (6.3) | 4,912 (6.3) | 491 (7.3) |  |
|  | Armed forces | 297 (0.3) | 279 (0.4) | 18 (0.3) |  |
|  | Arts, entertainment and recreation | 2,114 (2.5) | 1,994 (2.5) | 120 (1.8) |  |
|  | Other | 11,794 (13.8) | 10,976 (14.0) | 818 (12.1) |  |
|  | Unknown | 13,787 (16.2) | 12,625 (16.1) | 1,162 (17.2) |  |
| SOC Major Group, among<br>participants in employment (n, %) | Managers, directors and senior officials | 7,406 (8.7) | 6,872 (8.8) | 534 (7.9) | 19.1 |
|  | Professional occupations | 18,709 (21.9) | 17,331 (22.1) | 1,378 (20.4) |  |
|  | Associate professional and technical occupations | 13,770 (16.1) | 12,791 (16.3) | 979 (14.5) |  |
|  | Administrative and secretarial occupations | 9,802 (11.5) | 8,992 (11.5) | 810 (12.0) |  |
|  | Skilled trades occupations | 6,117 (7.2) | 5,680 (7.2) | 437 (6.5) |  |
|  | Caring, leisure and other service occupations | 5,375 (6.3) | 4,681 (6.0) | 694 (10.3) |  |
|  | Sales and customer service occupations | 4,296 (5.0) | 3,981 (5.1) | 315 (4.7) |  |
|  | Process, plant and machine operatives | 3,040 (3.6) | 2,826 (3.6) | 214 (3.2) |  |
|  | Elementary occupations | 4,116 (4.8) | 3,790 (4.8) | 326 (4.8) |  |
|  | Unknown | 12,640 (14.8) | 11,562 (14.7) | 1,078 (15.9) |  |
| Self-employment status, among<br>participants in employment (n, %) | Employee | 76,980 (90.3) | 70,776 (90.2) | 6,204 (91.7) | 5.4 |
|  | Self-employed | 8,291 (9.7) | 7,730 (9.8) | 561 (8.3) |  |

Notes: SD: standard deviation; SOC: Standard Occupational Classification. Area deprivation was based on the English Indices of Deprivation 2019, the Welsh Index of Multiple Deprivation 2019, the Scottish Index of Multiple Deprivation 2020, and the Northern Ireland Multiple Deprivation Measure 2017. Health conditions were self-reported rather than clinically diagnosed based on the survey question: "Do you have any physical or mental health conditions or illnesses lasting or expected to last 12 months or more (excluding any long-lasting COVID-19 symptoms)?"

**Supplementary Table 2.** Adjusted odds ratios for inactivity (excluding retirement) for participants currently reporting Long Covid compared with the pre-infection period, by time since SARS-CoV-2 infection and effect modifiers

| Effect modifier | Level | Time since infection | aOR (95% CI) | Uncorrected p-value | Corrected p-value |
| --- | --- | --- | --- | --- | --- |
| Age group (6 tests) | 16 to 49 years | 12 to <18 weeks | 0.73 (0.54 to 0.98) | Ref | Ref |
|  |  | 18 to <24 weeks | 0.93 (0.67 to 1.29) | Ref | Ref |
|  |  | 24 to <30 weeks | 0.98 (0.69 to 1.39) | Ref | Ref |
|  |  | 30 to <40 weeks | 1.18 (0.85 to 1.64) | Ref | Ref |
|  |  | 40 to <52 weeks | 0.99 (0.68 to 1.45) | Ref | Ref |
|  |  | ≥52 weeks | 1.01 (0.70 to 1.46) | Ref | Ref |
|  | 50 to 64 years | 12 to <18 weeks | 0.92 (0.71 to 1.18) | 0.24 | 0.59 |
|  |  | 18 to <24 weeks | 1.29 (0.97 to 1.72) | 0.13 | 0.49 |
|  |  | 24 to <30 weeks | 1.43 (1.04 to 1.95) | 0.11 | 0.49 |
|  |  | 30 to <40 weeks | 1.71 (1.28 to 2.29) | 0.09 | 0.49 |
|  |  | 40 to <52 weeks | 1.70 (1.22 to 2.37) | 0.03 | 0.49 |
|  |  | ≥52 weeks | 1.37 (1.02 to 1.83) | 0.21 | 0.59 |
| Sex (6 tests) | Male | 12 to <18 weeks | 0.99 (0.68 to 1.45) | Ref | Ref |
|  |  | 18 to <24 weeks | 0.90 (0.58 to 1.39) | Ref | Ref |
|  |  | 24 to <30 weeks | 1.03 (0.67 to 1.59) | Ref | Ref |
|  |  | 30 to <40 weeks | 1.57 (1.05 to 2.35) | Ref | Ref |
|  |  | 40 to <52 weeks | 1.41 (0.89 to 2.24) | Ref | Ref |
|  |  | ≥52 weeks | 0.80 (0.52 to 1.21) | Ref | Ref |
|  | Female | 12 to <18 weeks | 0.77 (0.62 to 0.96) | 0.25 | >0.99 |
|  |  | 18 to <24 weeks | 1.19 (0.93 to 1.54) | 0.27 | >0.99 |
|  |  | 24 to <30 weeks | 1.28 (0.97 to 1.69) | 0.41 | >0.99 |
|  |  | 30 to <40 weeks | 1.39 (1.08 to 1.81) | 0.63 | >0.99 |
|  |  | 40 to <52 weeks | 1.30 (0.97 to 1.75) | 0.77 | >0.99 |
|  |  | ≥52 weeks | 1.41 (1.07 to 1.86) | 0.03 | 0.37 |
| Ethnic group (6 tests) | Non-white | 12 to <18 weeks | 0.57 (0.27 to 1.20) | Ref | Ref |
|  |  | 18 to <24 weeks | 0.64 (0.28 to 1.49) | Ref | Ref |
|  |  | 24 to <30 weeks | 0.68 (0.28 to 1.69) | Ref | Ref |
|  |  | 30 to <40 weeks | 0.66 (0.28 to 1.57) | Ref | Ref |
|  |  | 40 to <52 weeks | 0.59 (0.25 to 1.42) | Ref | Ref |
|  |  | ≥52 weeks | 1.15 (0.55 to 2.41) | Ref | Ref |
|  | White | 12 to <18 weeks | 0.84 (0.69 to 1.03) | 0.32 | 0.95 |
|  |  | 18 to <24 weeks | 1.15 (0.92 to 1.44) | 0.19 | 0.77 |
|  |  | 24 to <30 weeks | 1.24 (0.98 to 1.58) | 0.21 | 0.77 |
|  |  | 30 to <40 weeks | 1.51 (1.21 to 1.90) | 0.07 | 0.51 |
|  |  | 40 to <52 weeks | 1.42 (1.10 to 1.85) | 0.06 | 0.51 |
|  |  | ≥52 weeks | 1.18 (0.93 to 1.51) | 0.95 | >0.99 |

**Supplementary Table 2 (continued)**
| Effect modifier | Level | Time since infection | aOR (95% CI) | Uncorrected p-value | Corrected p-value |
| --- | --- | --- | --- | --- | --- |
| Area deprivation quintile group (24 tests) | 1 (most deprived) | 12 to <18 weeks | 0.69 (0.47 to 1.02) | Ref | Ref |
|  |  | 18 to <24 weeks | 1.05 (0.68 to 1.62) | Ref | Ref |
|  |  | 24 to <30 weeks | 1.35 (0.83 to 2.19) | Ref | Ref |
|  |  | 30 to <40 weeks | 1.16 (0.76 to 1.78) | Ref | Ref |
|  |  | 40 to <52 weeks | 1.17 (0.72 to 1.90) | Ref | Ref |
|  |  | ≥52 weeks | 1.05 (0.69 to 1.59) | Ref | Ref |
|  | 2 | 12 to <18 weeks | 1.15 (0.77 to 1.71) | 0.07 | >0.99 |
|  |  | 18 to <24 weeks | 1.27 (0.83 to 1.96) | 0.53 | >0.99 |
|  |  | 24 to <30 weeks | 1.22 (0.75 to 1.98) | 0.78 | >0.99 |
|  |  | 30 to <40 weeks | 2.23 (1.44 to 3.47) | 0.04 | >0.99 |
|  |  | 40 to <52 weeks | 2.01 (1.23 to 3.28) | 0.13 | >0.99 |
|  |  | ≥52 weeks | 2.09 (1.31 to 3.32) | 0.03 | >0.99 |
|  | 3 | 12 to <18 weeks | 0.97 (0.61 to 1.55) | 0.27 | >0.99 |
|  |  | 18 to <24 weeks | 0.84 (0.49 to 1.46) | 0.54 | >0.99 |
|  |  | 24 to <30 weeks | 1.16 (0.66 to 2.03) | 0.69 | >0.99 |
|  |  | 30 to <40 weeks | 1.21 (0.70 to 2.10) | 0.91 | >0.99 |
|  |  | 40 to <52 weeks | 1.76 (0.92 to 3.36) | 0.32 | >0.99 |
|  |  | ≥52 weeks | 0.81 (0.45 to 1.46) | 0.48 | >0.99 |
|  | 4 | 12 to <18 weeks | 0.73 (0.47 to 1.13) | 0.88 | >0.99 |
|  |  | 18 to <24 weeks | 1.47 (0.88 to 2.47) | 0.32 | >0.99 |
|  |  | 24 to <30 weeks | 1.06 (0.61 to 1.84) | 0.52 | >0.99 |
|  |  | 30 to <40 weeks | 1.45 (0.87 to 2.42) | 0.52 | >0.99 |
|  |  | 40 to <52 weeks | 1.21 (0.65 to 2.23) | 0.94 | >0.99 |
|  |  | ≥52 weeks | 1.29 (0.72 to 2.32) | 0.57 | >0.99 |
|  | 5 (least deprived) | 12 to <18 weeks | 0.69 (0.42 to 1.12) | 0.97 | >0.99 |
|  |  | 18 to <24 weeks | 1.06 (0.61 to 1.84) | 0.97 | >0.99 |
|  |  | 24 to <30 weeks | 1.25 (0.72 to 2.20) | 0.85 | >0.99 |
|  |  | 30 to <40 weeks | 1.40 (0.81 to 2.44) | 0.60 | >0.99 |
|  |  | 40 to <52 weeks | 0.81 (0.41 to 1.57) | 0.37 | >0.99 |
|  |  | ≥52 weeks | 0.87 (0.44 to 1.70) | 0.64 | >0.99 |

**Supplementary Table 2 (continued)**
| Effect modifier | Level | Time since infection | aOR (95% CI) | Uncorrected p-value | Corrected p-value |
| --- | --- | --- | --- | --- | --- |
| Self-reported health/disability status (18 tests) | No long-term health conditions | 12 to <18 weeks | 0.71 (0.55 to 0.91) | Ref | Ref |
|  |  | 18 to <24 weeks | 1.13 (0.85 to 1.49) | Ref | Ref |
|  |  | 24 to <30 weeks | 0.98 (0.72 to 1.33) | Ref | Ref |
|  |  | 30 to <40 weeks | 1.42 (1.07 to 1.89) | Ref | Ref |
|  |  | 40 to <52 weeks | 1.09 (0.79 to 1.50) | Ref | Ref |
|  |  | ≥52 weeks | 1.13 (0.84 to 1.53) | Ref | Ref |
|  | Health conditions without impact to day-to-day activities | 12 to <18 weeks | 0.77 (0.42 to 1.44) | 0.79 | >0.99 |
|  |  | 18 to <24 weeks | 1.05 (0.53 to 2.09) | 0.86 | >0.99 |
|  |  | 24 to <30 weeks | 1.34 (0.66 to 2.73) | 0.42 | >0.99 |
|  |  | 30 to <40 weeks | 1.53 (0.78 to 3.00) | 0.84 | >0.99 |
|  |  | 40 to <52 weeks | 2.56 (1.34 to 4.91) | 0.02 | 0.66 |
|  |  | ≥52 weeks | 1.49 (0.77 to 2.87) | 0.45 | >0.99 |
|  | Day-to-day activities limited a little by health conditions | 12 to <18 weeks | 1.08 (0.66 to 1.78) | 0.14 | >0.99 |
|  |  | 18 to <24 weeks | 1.04 (0.59 to 1.84) | 0.81 | >0.99 |
|  |  | 24 to <30 weeks | 2.35 (1.20 to 4.63) | 0.02 | 0.66 |
|  |  | 30 to <40 weeks | 1.73 (0.91 to 3.26) | 0.59 | >0.99 |
|  |  | 40 to <52 weeks | 1.63 (0.78 to 3.41) | 0.33 | >0.99 |
|  |  | ≥52 weeks | 2.07 (1.09 to 3.93) | 0.10 | >0.99 |
|  | Day-to-day activities limited a lot by health conditions | 12 to <18 weeks | 1.15 (0.71 to 1.87) | 0.08 | >0.99 |
|  |  | 18 to <24 weeks | 1.24 (0.70 to 2.20) | 0.77 | >0.99 |
|  |  | 24 to <30 weeks | 1.38 (0.77 to 2.48) | 0.31 | >0.99 |
|  |  | 30 to <40 weeks | 1.35 (0.80 to 2.28) | 0.87 | >0.99 |
|  |  | 40 to <52 weeks | 1.42 (0.72 to 2.77) | 0.49 | >0.99 |
|  |  | ≥52 weeks | 0.72 (0.40 to 1.31) | 0.19 | >0.99 |
| Reinfected with SARS-CoV-2 (6 tests) | No | 12 to <18 weeks | 0.81 (0.67 to 0.98) | Ref | Ref |
|  |  | 18 to <24 weeks | 1.11 (0.89 to 1.39) | Ref | Ref |
|  |  | 24 to <30 weeks | 1.16 (0.91 to 1.47) | Ref | Ref |
|  |  | 30 to <40 weeks | 1.49 (1.18 to 1.87) | Ref | Ref |
|  |  | 40 to <52 weeks | 1.37 (1.04 to 1.81) | Ref | Ref |
|  |  | ≥52 weeks | 1.13 (0.86 to 1.47) | Ref | Ref |
|  | Yes | 12 to <18 weeks | 3.47 (0.77 to 15.64) | 0.06 | 0.83 |
|  |  | 18 to <24 weeks | 1.46 (0.55 to 3.88) | 0.59 | >0.99 |
|  |  | 24 to <30 weeks | 2.54 (0.98 to 6.62) | 0.12 | 0.83 |
|  |  | 30 to <40 weeks | 1.49 (0.72 to 3.07) | >0.99 | >0.99 |
|  |  | 40 to <52 weeks | 1.47 (0.75 to 2.88) | 0.85 | >0.99 |
|  |  | ≥52 weeks | 1.69 (0.99 to 2.90) | 0.17 | 0.83 |

**Supplementary Table 2 (continued)**
| Effect modifier | Level | Time since infection | aOR (95% CI) | Uncorrected p-value | Corrected p-value |
| --- | --- | --- | --- | --- | --- |
| Mode of data collection (6 tests) | Face-to-face | 12 to <18 weeks | 0.88 (0.72 to 1.08) | Ref | Ref |
|  |  | 18 to <24 weeks | 1.15 (0.90 to 1.46) | Ref | Ref |
|  |  | 24 to <30 weeks | 1.16 (0.89 to 1.52) | Ref | Ref |
|  |  | 30 to <40 weeks | 1.21 (0.93 to 1.56) | Ref | Ref |
|  |  | 40 to <52 weeks | 1.31 (0.97 to 1.77) | Ref | Ref |
|  |  | ≥52 weeks | 1.11 (0.84 to 1.46) | Ref | Ref |
|  | Remote | 12 to <18 weeks | 0.58 (0.31 to 1.07) | 0.20 | >0.99 |
|  |  | 18 to <24 weeks | 1.02 (0.62 to 1.69) | 0.69 | >0.99 |
|  |  | 24 to <30 weeks | 1.30 (0.79 to 2.12) | 0.70 | >0.99 |
|  |  | 30 to <40 weeks | 2.21 (1.46 to 3.35) | 0.01 | 0.22 |
|  |  | 40 to <52 weeks | 1.20 (0.76 to 1.87) | 0.73 | >0.99 |
|  |  | ≥52 weeks | 1.27 (0.90 to 1.79) | 0.52 | >0.99 |

Notes: aOR: adjusted odds ratio; CI: confidence interval; Ref: reference category. Estimates are from conditional logit models including the exposure variable interacted with each of the effect modifiers, adjusted for calendar day of study assessment, current age, and interactions between calendar day and each of current age, sex, self-reported health/disability status at survey enrolment, and each of the effect modifiers (excluding reinfection status and data collection mode). P-values were corrected using the Benjamini-Yekutieli method.

**Supplementary Table 3.** Adjusted odds ratios for long-term (≥4 weeks) absence for participants currently reporting Long Covid compared with the pre-infection period, by time since SARS-CoV-2 infection and effect modifiers

| Effect modifier | Level | Time since infection | aOR (95% CI) | Uncorrected p-value | Corrected p-value |
| --- | --- | --- | --- | --- | --- |
| Age group (6 tests) | 16 to 49 years | 12 to <18 weeks | 0.99 (0.65 to 1.52) | Ref | Ref |
|  |  | 18 to <24 weeks | 1.36 (0.89 to 2.09) | Ref | Ref |
|  |  | 24 to <30 weeks | 1.05 (0.64 to 1.70) | Ref | Ref |
|  |  | 30 to <40 weeks | 0.85 (0.52 to 1.40) | Ref | Ref |
|  |  | 40 to <52 weeks | 0.84 (0.48 to 1.48) | Ref | Ref |
|  |  | ≥52 weeks | 0.51 (0.27 to 0.97) | Ref | Ref |
|  | 50 to 64 years | 12 to <18 weeks | 1.46 (0.97 to 2.20) | 0.20 | 0.83 |
|  |  | 18 to <24 weeks | 1.48 (0.97 to 2.28) | 0.78 | >0.99 |
|  |  | 24 to <30 weeks | 2.03 (1.27 to 3.26) | 0.053 | 0.39 |
|  |  | 30 to <40 weeks | 1.28 (0.82 to 2.01) | 0.23 | 0.83 |
|  |  | 40 to <52 weeks | 1.00 (0.61 to 1.66) | 0.64 | >0.99 |
|  |  | ≥52 weeks | 1.26 (0.73 to 2.18) | 0.03 | 0.39 |
| Sex (6 tests) | Male | 12 to <18 weeks | 1.32 (0.79 to 2.23) | Ref | Ref |
|  |  | 18 to <24 weeks | 1.33 (0.77 to 2.31) | Ref | Ref |
|  |  | 24 to <30 weeks | 1.88 (1.05 to 3.39) | Ref | Ref |
|  |  | 30 to <40 weeks | 1.13 (0.62 to 2.09) | Ref | Ref |
|  |  | 40 to <52 weeks | 0.93 (0.46 to 1.89) | Ref | Ref |
|  |  | ≥52 weeks | 0.67 (0.31 to 1.44) | Ref | Ref |
|  | Female | 12 to <18 weeks | 1.15 (0.81 to 1.65) | 0.67 | >0.99 |
|  |  | 18 to <24 weeks | 1.45 (1.01 to 2.09) | 0.80 | >0.99 |
|  |  | 24 to <30 weeks | 1.28 (0.85 to 1.95) | 0.30 | >0.99 |
|  |  | 30 to <40 weeks | 1.00 (0.67 to 1.48) | 0.73 | >0.99 |
|  |  | 40 to <52 weeks | 0.86 (0.55 to 1.35) | 0.84 | >0.99 |
|  |  | ≥52 weeks | 0.92 (0.56 to 1.53) | 0.49 | >0.99 |
| Ethnic group (6 tests) | Non-white | 12 to <18 weeks | 1.26 (0.31 to 5.06) | Ref | Ref |
|  |  | 18 to <24 weeks | 1.08 (0.24 to 4.91) | Ref | Ref |
|  |  | 24 to <30 weeks | 0.51 (0.06 to 4.23) | Ref | Ref |
|  |  | 30 to <40 weeks | 0.82 (0.19 to 3.57) | Ref | Ref |
|  |  | 40 to <52 weeks | 0.32 (0.05 to 1.98) | Ref | Ref |
|  |  | ≥52 weeks | 0.27 (0.05 to 1.40) | Ref | Ref |
|  | White | 12 to <18 weeks | 1.20 (0.89 to 1.63) | 0.95 | >0.99 |
|  |  | 18 to <24 weeks | 1.41 (1.04 to 1.93) | 0.74 | >0.99 |
|  |  | 24 to <30 weeks | 1.52 (1.07 to 2.14) | 0.32 | >0.99 |
|  |  | 30 to <40 weeks | 1.05 (0.75 to 1.48) | 0.75 | >0.99 |
|  |  | 40 to <52 weeks | 0.93 (0.63 to 1.38) | 0.26 | >0.99 |
|  |  | ≥52 weeks | 0.89 (0.58 to 1.38) | 0.17 | >0.99 |

**Supplementary Table 3 (continued)**
| Effect modifier | Level | Time since infection | aOR (95% CI) | Uncorrected p-value | Corrected p-value |
| --- | --- | --- | --- | --- | --- |
| Area deprivation quintile group (24 tests) | 1 (most deprived) | 12 to <18 weeks | 0.71 (0.33 to 1.50) | Ref | Ref |
|  |  | 18 to <24 weeks | 0.41 (0.16 to 1.08) | Ref | Ref |
|  |  | 24 to <30 weeks | 0.85 (0.35 to 2.05) | Ref | Ref |
|  |  | 30 to <40 weeks | 0.72 (0.29 to 1.80) | Ref | Ref |
|  |  | 40 to <52 weeks | 0.73 (0.29 to 1.84) | Ref | Ref |
|  |  | ≥52 weeks | 0.43 (0.15 to 1.20) | Ref | Ref |
|  | 2 | 12 to <18 weeks | 1.29 (0.66 to 2.50) | 0.24 | >0.99 |
|  |  | 18 to <24 weeks | 2.26 (1.25 to 4.09) | 0.003 | 0.26 |
|  |  | 24 to <30 weeks | 1.63 (0.81 to 3.28) | 0.25 | >0.99 |
|  |  | 30 to <40 weeks | 1.10 (0.53 to 2.27) | 0.49 | >0.99 |
|  |  | 40 to <52 weeks | 1.30 (0.56 to 2.98) | 0.37 | >0.99 |
|  |  | ≥52 weeks | 1.58 (0.67 to 3.73) | 0.06 | >0.99 |
|  | 3 | 12 to <18 weeks | 1.73 (0.91 to 3.32) | 0.08 | >0.99 |
|  |  | 18 to <24 weeks | 1.54 (0.78 to 3.03) | 0.03 | 0.85 |
|  |  | 24 to <30 weeks | 1.87 (0.86 to 4.02) | 0.19 | >0.99 |
|  |  | 30 to <40 weeks | 1.32 (0.64 to 2.76) | 0.31 | >0.99 |
|  |  | 40 to <52 weeks | 0.74 (0.30 to 1.84) | 0.99 | >0.99 |
|  |  | ≥52 weeks | 0.33 (0.11 to 1.06) | 0.75 | >0.99 |
|  | 4 | 12 to <18 weeks | 1.55 (0.81 to 2.99) | 0.12 | >0.99 |
|  |  | 18 to <24 weeks | 2.13 (1.09 to 4.16) | 0.01 | 0.26 |
|  |  | 24 to <30 weeks | 2.34 (1.15 to 4.75) | 0.08 | >0.99 |
|  |  | 30 to <40 weeks | 1.18 (0.58 to 2.42) | 0.40 | >0.99 |
|  |  | 40 to <52 weeks | 0.83 (0.37 to 1.87) | 0.85 | >0.99 |
|  |  | ≥52 weeks | 0.80 (0.33 to 1.98) | 0.37 | >0.99 |
|  | 5 (least deprived) | 12 to <18 weeks | 0.96 (0.51 to 1.81) | 0.54 | >0.99 |
|  |  | 18 to <24 weeks | 1.09 (0.55 to 2.13) | 0.11 | >0.99 |
|  |  | 24 to <30 weeks | 0.95 (0.42 to 2.14) | 0.85 | >0.99 |
|  |  | 30 to <40 weeks | 0.88 (0.44 to 1.79) | 0.73 | >0.99 |
|  |  | 40 to <52 weeks | 1.04 (0.45 to 2.39) | 0.58 | >0.99 |
|  |  | ≥52 weeks | 1.45 (0.56 to 3.74) | 0.09 | >0.99 |

**Supplementary Table 3 (continued)**
| Effect modifier | Level | Time since infection | aOR (95% CI) | Uncorrected p-value | Corrected p-value |
| --- | --- | --- | --- | --- | --- |
| Presence of self-reported health conditions (6 tests) | No | 12 to <18 weeks | 1.26 (0.89 to 1.79) | Ref | Ref |
|  |  | 18 to <24 weeks | 1.96 (1.38 to 2.78) | Ref | Ref |
|  |  | 24 to <30 weeks | 1.42 (0.95 to 2.12) | Ref | Ref |
|  |  | 30 to <40 weeks | 1.00 (0.67 to 1.49) | Ref | Ref |
|  |  | 40 to <52 weeks | 0.97 (0.62 to 1.53) | Ref | Ref |
|  |  | ≥52 weeks | 1.05 (0.65 to 1.71) | Ref | Ref |
|  | Yes | 12 to <18 weeks | 1.06 (0.61 to 1.85) | 0.61 | >0.99 |
|  |  | 18 to <24 weeks | 0.58 (0.31 to 1.10) | 0.001 | 0.02 |
|  |  | 24 to <30 weeks | 1.51 (0.80 to 2.86) | 0.87 | >0.99 |
|  |  | 30 to <40 weeks | 1.04 (0.57 to 1.90) | 0.91 | >0.99 |
|  |  | 40 to <52 weeks | 0.67 (0.33 to 1.37) | 0.40 | >0.99 |
|  |  | ≥52 weeks | 0.45 (0.20 to 1.01) | 0.08 | 0.57 |
| Reinfected with SARS-CoV-2 (6 tests) | No | 12 to <18 weeks | 1.17 (0.87 to 1.58) | Ref | Ref |
|  |  | 18 to <24 weeks | 1.35 (0.98 to 1.85) | Ref | Ref |
|  |  | 24 to <30 weeks | 1.55 (1.08 to 2.22) | Ref | Ref |
|  |  | 30 to <40 weeks | 0.93 (0.65 to 1.34) | Ref | Ref |
|  |  | 40 to <52 weeks | 0.85 (0.56 to 1.30) | Ref | Ref |
|  |  | ≥52 weeks | 0.69 (0.42 to 1.11) | Ref | Ref |
|  | Yes | 12 to <18 weeks | 4.46 (0.51 to 39.38) | 0.23 | >0.99 |
|  |  | 18 to <24 weeks | 1.93 (0.64 to 5.83) | 0.53 | >0.99 |
|  |  | 24 to <30 weeks | 0.74 (0.24 to 2.28) | 0.22 | >0.99 |
|  |  | 30 to <40 weeks | 1.33 (0.55 to 3.22) | 0.46 | >0.99 |
|  |  | 40 to <52 weeks | 0.71 (0.28 to 1.84) | 0.73 | >0.99 |
|  |  | ≥52 weeks | 0.96 (0.44 to 2.10) | 0.40 | >0.99 |
| Mode of data collection (6 tests) | Face-to-face | 12 to <18 weeks | 1.00 (0.71 to 1.41) | Ref | Ref |
|  |  | 18 to <24 weeks | 1.01 (0.69 to 1.49) | Ref | Ref |
|  |  | 24 to <30 weeks | 1.15 (0.72 to 1.81) | Ref | Ref |
|  |  | 30 to <40 weeks | 0.89 (0.56 to 1.41) | Ref | Ref |
|  |  | 40 to <52 weeks | 0.95 (0.57 to 1.59) | Ref | Ref |
|  |  | ≥52 weeks | 0.72 (0.42 to 1.23) | Ref | Ref |
|  | Remote | 12 to <18 weeks | 2.19 (1.14 to 4.21) | 0.04 | 0.19 |
|  |  | 18 to <24 weeks | 2.37 (1.41 to 3.99) | 0.01 | 0.13 |
|  |  | 24 to <30 weeks | 1.79 (1.07 to 2.98) | 0.20 | 0.73 |
|  |  | 30 to <40 weeks | 1.05 (0.66 to 1.69) | 0.61 | >0.99 |
|  |  | 40 to <52 weeks | 0.44 (0.25 to 0.79) | 0.04 | 0.19 |
|  |  | ≥52 weeks | 0.63 (0.37 to 1.07) | 0.59 | >0.99 |

**Supplementary Table 3 (continued)**
| Effect modifier | Level | Time since infection | aOR (95% CI) | Uncorrected p-value | Corrected p-value |
| --- | --- | --- | --- | --- | --- |
| Employment sector, among participants in employment (42 tests) | Teaching and education | 12 to <18 weeks | 0.89 (0.43 to 1.81) | Ref | Ref |
|  |  | 18 to <24 weeks | 1.12 (0.59 to 2.11) | Ref | Ref |
|  |  | 24 to <30 weeks | 2.06 (1.08 to 3.96) | Ref | Ref |
|  |  | 30 to <40 weeks | 1.00 (0.52 to 1.90) | Ref | Ref |
|  |  | 40 to <52 weeks | 0.69 (0.31 to 1.56) | Ref | Ref |
|  |  | ≥52 weeks | 1.39 (0.56 to 3.44) | Ref | Ref |
|  | Health or social care | 12 to <18 weeks | 2.07 (1.15 to 3.73) | 0.07 | >0.99 |
|  |  | 18 to <24 weeks | 2.00 (1.04 to 3.84) | 0.21 | >0.99 |
|  |  | 24 to <30 weeks | 0.99 (0.42 to 2.31) | 0.18 | >0.99 |
|  |  | 30 to <40 weeks | 0.82 (0.38 to 1.78) | 0.71 | >0.99 |
|  |  | 40 to <52 weeks | 0.85 (0.35 to 2.04) | 0.74 | >0.99 |
|  |  | ≥52 weeks | 0.46 (0.16 to 1.31) | 0.12 | >0.99 |
|  | Transport | 12 to <18 weeks | 0.23 (0.02 to 2.19) | 0.26 | >0.99 |
|  |  | 18 to <24 weeks | 0.77 (0.11 to 5.17) | 0.72 | >0.99 |
|  |  | 24 to <30 weeks | 0.53 (0.10 to 2.70) | 0.13 | >0.99 |
|  |  | 30 to <40 weeks | 0.52 (0.10 to 2.71) | 0.47 | >0.99 |
|  |  | 40 to <52 weeks | 0.34 (0.05 to 2.34) | 0.51 | >0.99 |
|  |  | ≥52 weeks | 1.48 (0.25 to 8.68) | 0.95 | >0.99 |
|  | Retail and wholesale | 12 to <18 weeks | 1.30 (0.30 to 5.63) | 0.64 | >0.99 |
|  |  | 18 to <24 weeks | 0.49 (0.09 to 2.78) | 0.38 | >0.99 |
|  |  | 24 to <30 weeks | 2.05 (0.39 to 10.82) | 0.99 | >0.99 |
|  |  | 30 to <40 weeks | 0.96 (0.22 to 4.11) | 0.96 | >0.99 |
|  |  | 40 to <52 weeks | 0.97 (0.23 to 4.12) | 0.69 | >0.99 |
|  |  | ≥52 weeks | 0.74 (0.14 to 3.81) | 0.51 | >0.99 |
|  | Hospitality | 12 to <18 weeks | 0.65 (0.07 to 5.85) | 0.80 | >0.99 |
|  |  | 18 to <24 weeks | 3.35 (0.47 to 23.84) | 0.30 | >0.99 |
|  |  | 24 to <30 weeks | 6.69 (0.84 to 53.53) | 0.29 | >0.99 |
|  |  | 30 to <40 weeks | 2.98 (0.39 to 22.90) | 0.32 | >0.99 |
|  |  | 40 to <52 weeks | 6.99 (0.85 to 57.43) | 0.04 | >0.99 |
|  |  | ≥52 weeks | 3.97 (0.25 to 62.24) | 0.48 | >0.99 |
|  | Manufacturing and construction | 12 to <18 weeks | 1.30 (0.42 to 4.05) | 0.58 | >0.99 |
|  |  | 18 to <24 weeks | 2.06 (0.64 to 6.61) | 0.36 | >0.99 |
|  |  | 24 to <30 weeks | 2.84 (0.78 to 10.40) | 0.67 | >0.99 |
|  |  | 30 to <40 weeks | 1.26 (0.21 to 7.63) | 0.81 | >0.99 |
|  |  | 40 to <52 weeks | 2.22 (0.35 to 13.91) | 0.26 | >0.99 |
|  |  | ≥52 weeks | 1.12 (0.17 to 7.18) | 0.84 | >0.99 |

**Supplementary Table 3 (continued)**
| Effect modifier | Level | Time since infection | aOR (95% CI) | Uncorrected p-value | Corrected p-value |
| --- | --- | --- | --- | --- | --- |
| Employment sector, among participants in employment (42 tests) (continued) | Civil service and local government | 12 to <18 weeks | 0.92 (0.32 to 2.68) | 0.95 | >0.99 |
|  |  | 18 to <24 weeks | 0.44 (0.12 to 1.55) | 0.19 | >0.99 |
|  |  | 24 to <30 weeks | 0.28 (0.05 to 1.47) | 0.03 | >0.99 |
|  |  | 30 to <40 weeks | 0.40 (0.10 to 1.51) | 0.22 | >0.99 |
|  |  | 40 to <52 weeks | 0.11 (0.02 to 0.62) | 0.06 | >0.99 |
|  |  | ≥52 weeks | 0.06 (0.01 to 0.37) | 0.002 | 0.41 |
|  | Other | 12 to <18 weeks | 0.72 (0.34 to 1.52) | 0.69 | >0.99 |
|  |  | 18 to <24 weeks | 1.53 (0.78 to 3.00) | 0.50 | >0.99 |
|  |  | 24 to <30 weeks | 1.17 (0.50 to 2.70) | 0.29 | >0.99 |
|  |  | 30 to <40 weeks | 0.93 (0.40 to 2.15) | 0.89 | >0.99 |
|  |  | 40 to <52 weeks | 0.23 (0.08 to 0.69) | 0.11 | >0.99 |
|  |  | ≥52 weeks | 0.27 (0.10 to 0.72) | 0.01 | >0.99 |
| SOC Major Group, among participants in employment (42 tests) | Managers, directors and senior officials | 12 to <18 weeks | 1.16 (0.32 to 4.19) | Ref | Ref |
|  |  | 18 to <24 weeks | 1.56 (0.32 to 7.65) | Ref | Ref |
|  |  | 24 to <30 weeks | 3.02 (0.79 to 11.45) | Ref | Ref |
|  |  | 30 to <40 weeks | 1.09 (0.20 to 5.91) | Ref | Ref |
|  |  | 40 to <52 weeks | 2.41 (0.47 to 12.30) | Ref | Ref |
|  |  | ≥52 weeks | 0.57 (0.07 to 4.67) | Ref | Ref |
|  | Professional occupations | 12 to <18 weeks | 1.35 (0.74 to 2.47) | 0.83 | >0.99 |
|  |  | 18 to <24 weeks | 1.68 (0.88 to 3.19) | 0.94 | >0.99 |
|  |  | 24 to <30 weeks | 1.35 (0.66 to 2.77) | 0.30 | >0.99 |
|  |  | 30 to <40 weeks | 1.12 (0.55 to 2.25) | 0.98 | >0.99 |
|  |  | 40 to <52 weeks | 0.87 (0.40 to 1.92) | 0.27 | >0.99 |
|  |  | ≥52 weeks | 0.84 (0.35 to 2.02) | 0.73 | >0.99 |
|  | Associate professional and technical occupations | 12 to <18 weeks | 1.31 (0.50 to 3.39) | 0.88 | >0.99 |
|  |  | 18 to <24 weeks | 1.72 (0.74 to 4.00) | 0.92 | >0.99 |
|  |  | 24 to <30 weeks | 1.58 (0.46 to 5.49) | 0.49 | >0.99 |
|  |  | 30 to <40 weeks | 1.73 (0.61 to 4.92) | 0.65 | >0.99 |
|  |  | 40 to <52 weeks | 0.74 (0.21 to 2.57) | 0.26 | >0.99 |
|  |  | ≥52 weeks | 0.70 (0.18 to 2.69) | 0.87 | >0.99 |
|  | Administrative and secretarial occupations | 12 to <18 weeks | 0.60 (0.18 to 2.05) | 0.47 | >0.99 |
|  |  | 18 to <24 weeks | 1.13 (0.41 to 3.08) | 0.73 | >0.99 |
|  |  | 24 to <30 weeks | 1.72 (0.50 to 5.85) | 0.54 | >0.99 |
|  |  | 30 to <40 weeks | 1.36 (0.46 to 4.04) | 0.83 | >0.99 |
|  |  | 40 to <52 weeks | 0.46 (0.11 to 1.86) | 0.13 | >0.99 |
|  |  | ≥52 weeks | 0.73 (0.20 to 2.60) | 0.84 | >0.99 |

**Supplementary Table 3 (continued)**
| Effect modifier | Level | Time since infection | aOR (95% CI) | Uncorrected p-value | Corrected p-value |
| --- | --- | --- | --- | --- | --- |
| SOC Major Group, among participants in employment (42 tests)<br>(continued) | Skilled trades occupations | 12 to <18 weeks | 0.12 (0.01 to 1.43) | 0.11 | >0.99 |
|  |  | 18 to <24 weeks | 0.67 (0.16 to 2.75) | 0.43 | >0.99 |
|  |  | 24 to <30 weeks | 3.69 (0.87 to 15.69) | 0.84 | >0.99 |
|  |  | 30 to <40 weeks | 0.45 (0.08 to 2.49) | 0.47 | >0.99 |
|  |  | 40 to <52 weeks | 1.70 (0.43 to 6.70) | 0.74 | >0.99 |
|  |  | ≥52 weeks | 1.13 (0.22 to 5.91) | 0.61 | >0.99 |
|  | Caring, leisure and other service occupations | 12 to <18 weeks | 1.67 (0.82 to 3.41) | 0.62 | >0.99 |
|  |  | 18 to <24 weeks | 1.28 (0.56 to 2.93) | 0.83 | >0.99 |
|  |  | 24 to <30 weeks | 1.76 (0.74 to 4.15) | 0.50 | >0.99 |
|  |  | 30 to <40 weeks | 1.55 (0.74 to 3.27) | 0.71 | >0.99 |
|  |  | 40 to <52 weeks | 0.70 (0.27 to 1.83) | 0.20 | >0.99 |
|  |  | ≥52 weeks | 0.59 (0.20 to 1.75) | 0.97 | >0.99 |
|  | Sales and customer service occupations | 12 to <18 weeks | 1.69 (0.33 to 8.79) | 0.72 | >0.99 |
|  |  | 18 to <24 weeks | 3.51 (0.49 to 25.18) | 0.53 | >0.99 |
|  |  | 24 to <30 weeks | <0.01 (<0.01 to >99.9) | 0.98 | >0.99 |
|  |  | 30 to <40 weeks | 1.67 (0.35 to 8.08) | 0.72 | >0.99 |
|  |  | 40 to <52 weeks | 0.48 (0.10 to 2.39) | 0.17 | >0.99 |
|  |  | ≥52 weeks | 0.47 (0.11 to 2.03) | 0.89 | >0.99 |
|  | Process, plant and machine operatives; and elementary occupations | 12 to <18 weeks | 1.22 (0.47 to 3.15) | 0.95 | >0.99 |
|  |  | 18 to <24 weeks | 1.91 (0.68 to 5.36) | 0.84 | >0.99 |
|  |  | 24 to <30 weeks | 2.12 (0.78 to 5.77) | 0.68 | >0.99 |
|  |  | 30 to <40 weeks | 1.34 (0.42 to 4.29) | 0.84 | >0.99 |
|  |  | 40 to <52 weeks | 1.24 (0.23 to 6.59) | 0.58 | >0.99 |
|  |  | ≥52 weeks | 1.22 (0.24 to 6.23) | 0.57 | >0.99 |
| Self-employment status, among participants in employment (6 tests) | Employee | 12 to <18 weeks | 1.21 (0.88 to 1.67) | Ref | Ref |
|  |  | 18 to <24 weeks | 1.39 (1.00 to 1.92) | Ref | Ref |
|  |  | 24 to <30 weeks | 1.35 (0.94 to 1.94) | Ref | Ref |
|  |  | 30 to <40 weeks | 1.14 (0.80 to 1.62) | Ref | Ref |
|  |  | 40 to <52 weeks | 0.75 (0.50 to 1.13) | Ref | Ref |
|  |  | ≥52 weeks | 0.71 (0.45 to 1.11) | Ref | Ref |
|  | Self-employed | 12 to <18 weeks | 1.04 (0.45 to 2.40) | 0.74 | >0.99 |
|  |  | 18 to <24 weeks | 1.23 (0.51 to 2.97) | 0.80 | >0.99 |
|  |  | 24 to <30 weeks | 1.35 (0.49 to 3.73) | >0.99 | >0.99 |
|  |  | 30 to <40 weeks | 0.18 (0.05 to 0.68) | 0.01 | 0.12 |
|  |  | 40 to <52 weeks | 1.38 (0.53 to 3.63) | 0.24 | >0.99 |
|  |  | ≥52 weeks | 0.79 (0.28 to 2.21) | 0.84 | >0.99 |

Notes: aOR: adjusted odds ratio; CI: confidence interval; Ref: reference category. Estimates are from conditional logit models including the exposure variable interacted with each of the effect modifiers, adjusted for calendar day of study assessment, current age, and interactions between calendar day and each of current age, sex, self-reported health/disability status at survey enrolment, and each of the effect modifiers (excluding reinfection status and data collection mode). Models were fitted to study assessments from 1 October 2021 when participants were in employment. P-values were corrected using the Benjamini-Yekutieli method.

**Supplementary Table 4.** Number and percentage of households invited to participate in the COVID-19 Infection Survey who subsequently enrolled, by country and phase of study

| Study phase | England | Wales | Northern Ireland | Scotland |
| --- | --- | --- | --- | --- |
| Initial invitation | 10,266 (51%) | 7,031 (41%) | 7,373 (43%) | N/A |
| Extension period | 39,392 (43%) | N/A | N/A | N/A |
| AddressBase | 173,583 (12%) | 7,051 (14%) | N/A | 23,217 (13%) |

Notes: The initial invitation phase was open to previous respondents to ONS surveys who had consented to participate in future research, and started on 26 April 2020 in England, 29 June 2020 in Wales, and 26 July 2020 in Northern Ireland. The extension period refers to the period of time beyond the initial pilot phase of the study when the sample was increased, and started on 31 May 2020 in England. Sampling from AddressBase started on 13 July 2020 in England, 5 October 2020 in Wales, and 14 September 2020 in Scotland, and involved randomly selecting addresses from an address list. Enrolment rates are as of 31 January 2022, when recruitment into the study ended, and are taken from the technical dataset accompanying the official COVID-19 Infection Survey publication: https://www.ons.gov.uk/peoplepopulationandcommunity/healthandsocialcare/conditionsanddiseases/datasets/covid19infectionsurveytechnicaldata

**Supplementary Figure 1.**
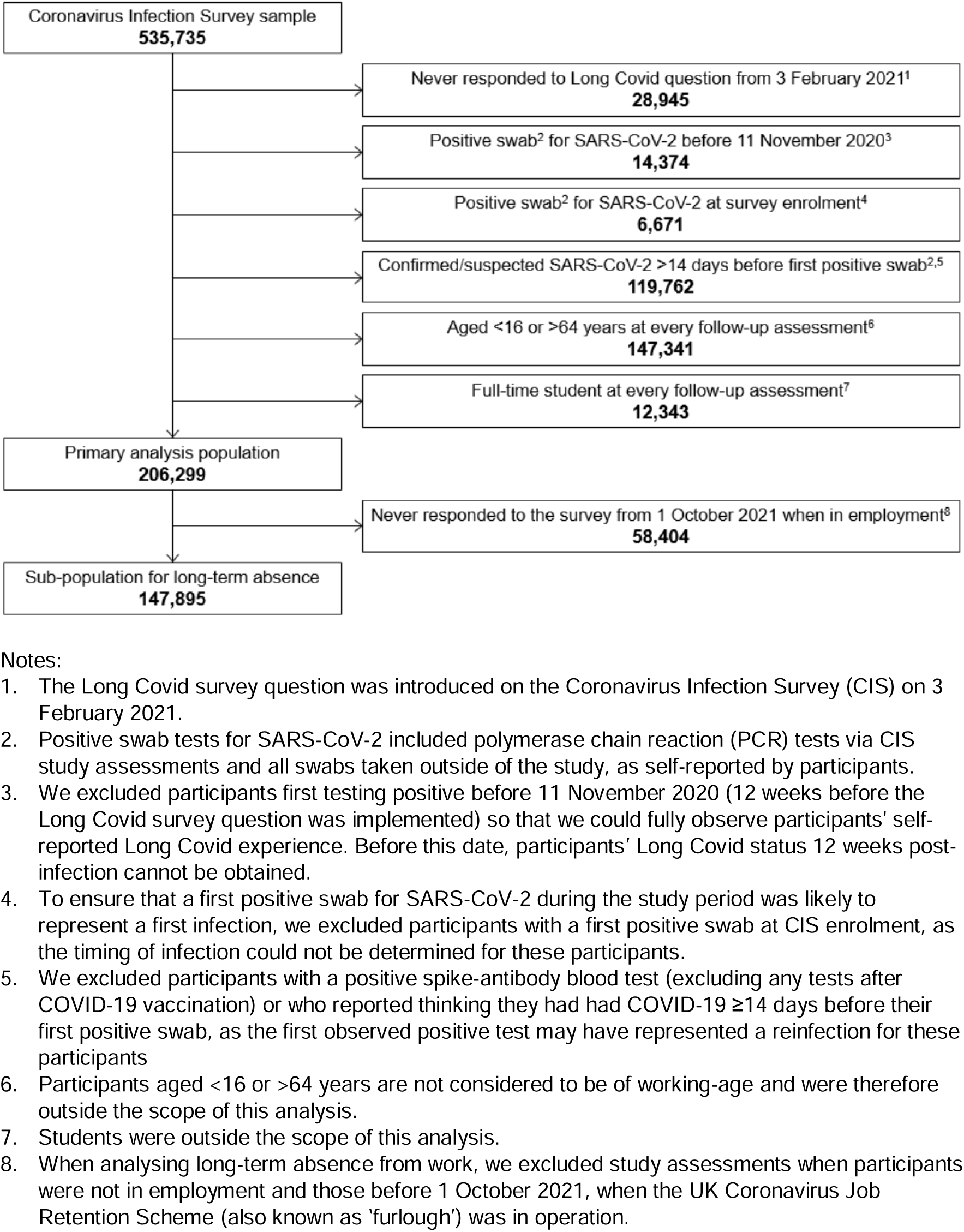
Study participant flow diagram for the analysis population

**Supplementary Figure 2.**
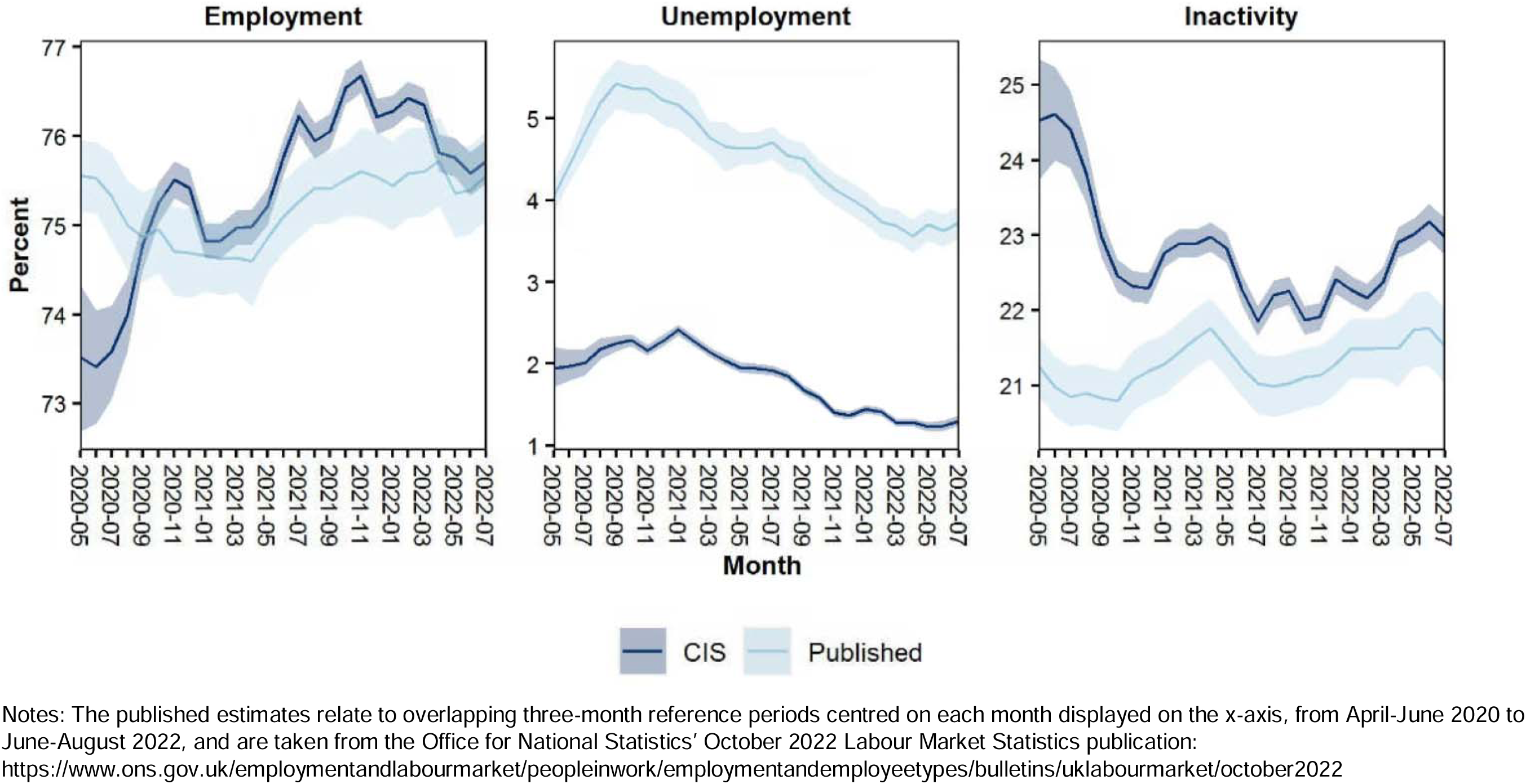
Rates of employment, unemployment and labour market inactivity among COVID-19 Infection Survey (CIS) participants aged 16 to 64 years (unweighted), compared with published national statistics from the UK Labour Force Survey

## SUPPLEMENTARY APPENDIX 1

### Sensitivity analysis for labour market inactivity

We performed several sensitivity analyses for the primary outcome, labour market inactivity:

- First, we restricted the analysis to participants who tested positive for SARS-CoV-2 during follow-up to mitigate against selection effects
- Second, we excluded study assessments when study participants were retired, and therefore ineligible to be otherwise inactive
- Third, we investigated alternative specifications of the restricted cubic spline for modelling calendar time and age

These analyses are illustrated in the following figures. All estimates and inferences are similar to those presented in the main analysis.

**Sensitivity analysis 1:**
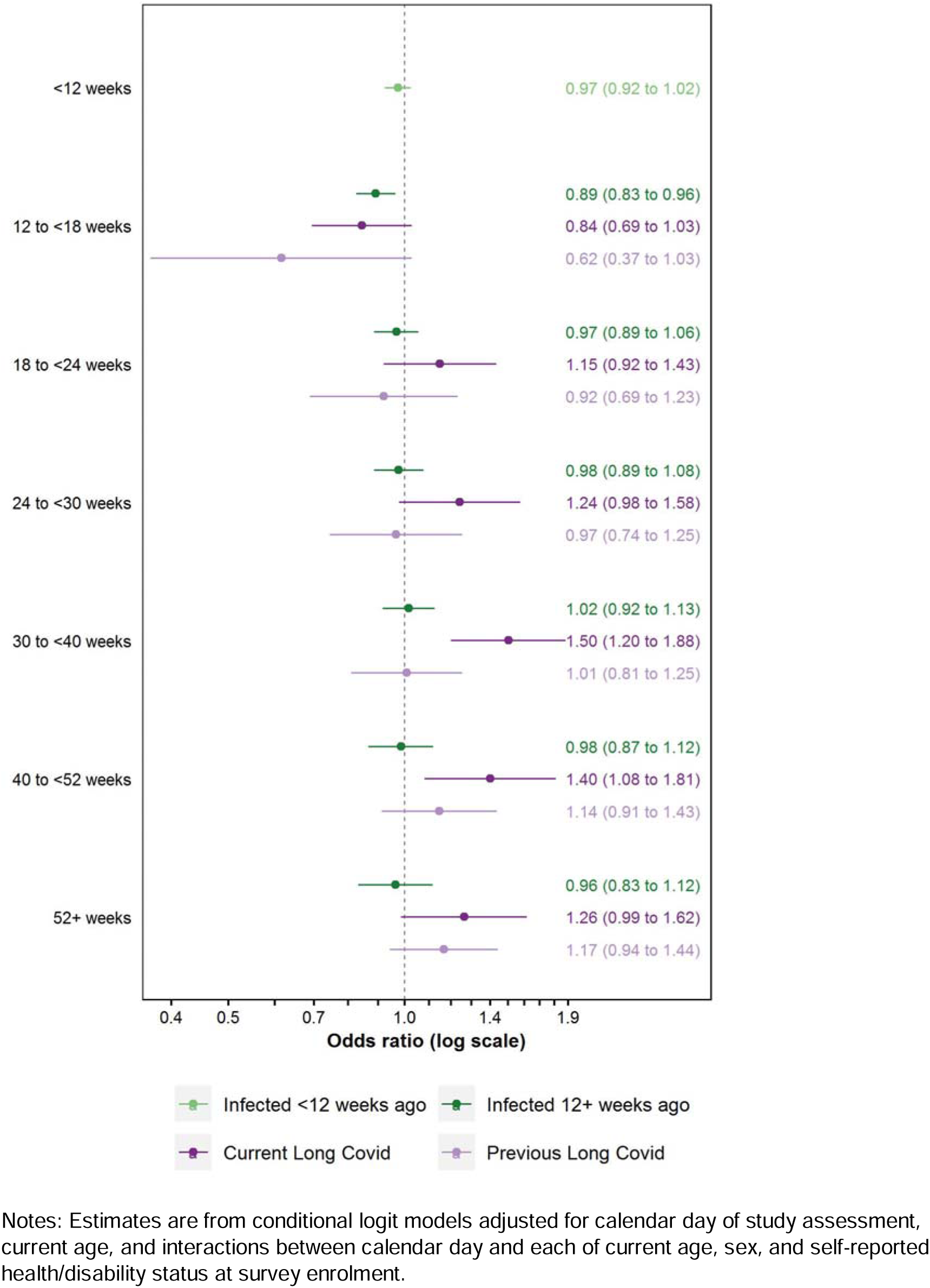
Adjusted odds ratios for inactivity (excluding retirement) compared with the pre-infection period, after restricting the analysis population to ever-infected participants

**Sensitivity analysis 2:**
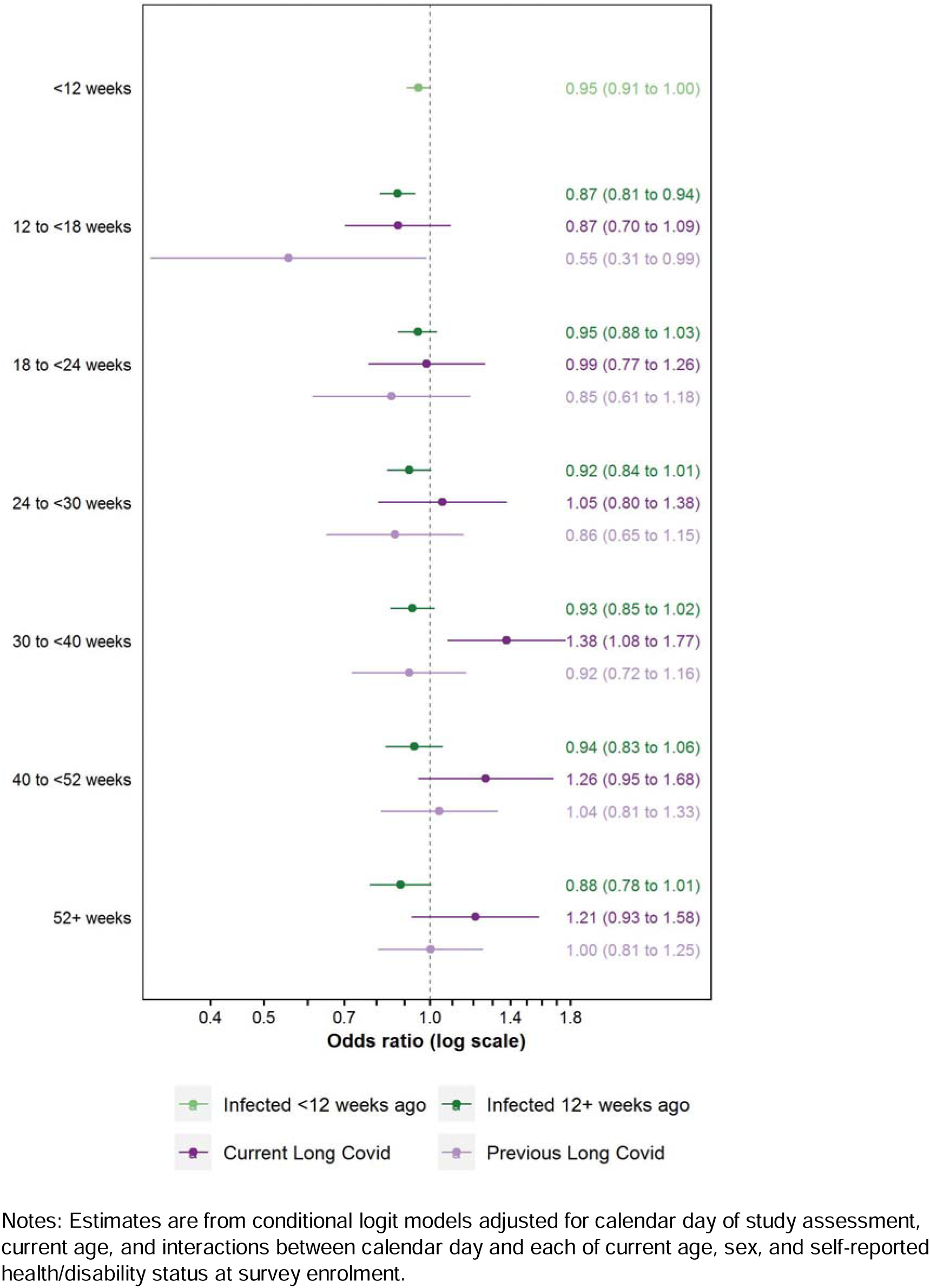
Adjusted odds ratios for inactivity (excluding retirement) compared with the pre-infection period, after excluding study assessments when participants were retired

**Sensitivity analysis 3a:**
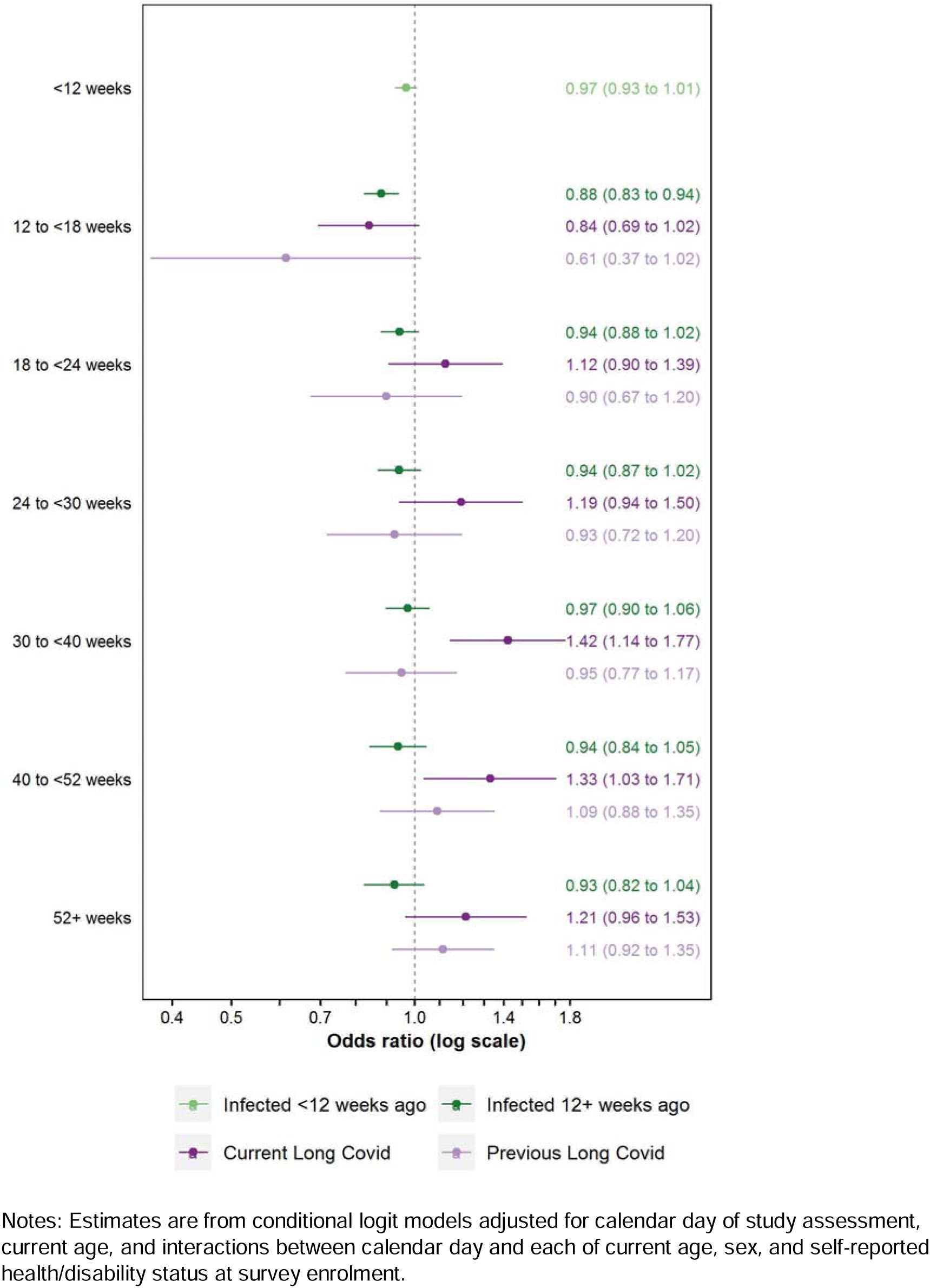
Adjusted odds ratios for inactivity (excluding retirement) compared with the pre-infection period, after increasing the number of internal knots in splines for calendar time and age from one to two

**Sensitivity analysis 3b:**
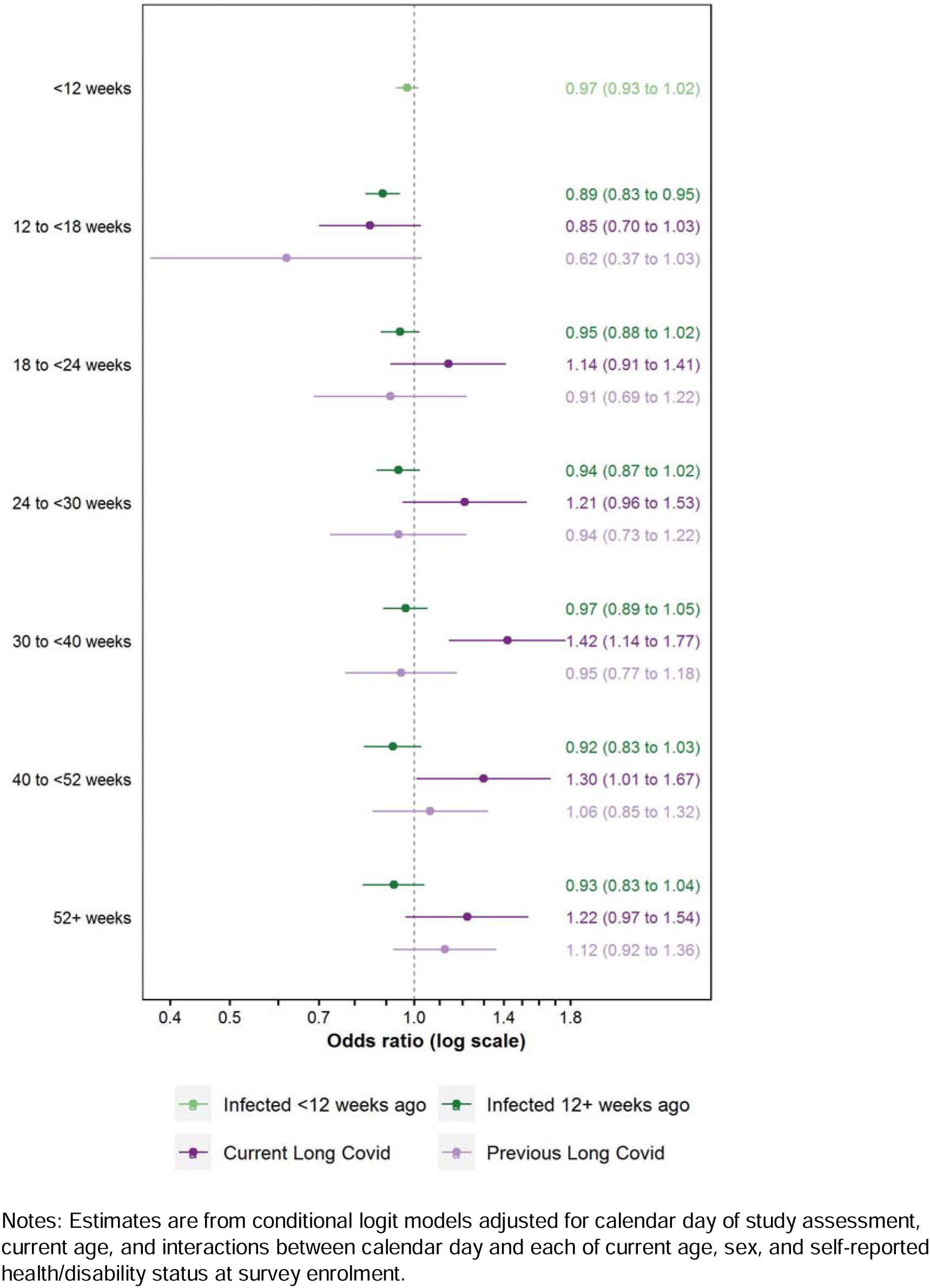
Adjusted odds ratios for inactivity (excluding retirement) compared with the pre-infection period, after increasing the number of internal knots in splines for calendar time and age from one to three

## SUPPLEMENTARY APPENDIX 2

### Methodology for estimating labour market inactivity attributable to Long Covid

#### Point estimates

1. For each time-since-infection stratum (*i*), use the published estimates of the total number of people reporting Long Covid (*n_i_*) and those who are inactive (*y_i_*)^1^ to calculate the probability of inactivity among people reporting Long Covid (*p_i_*): 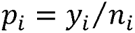 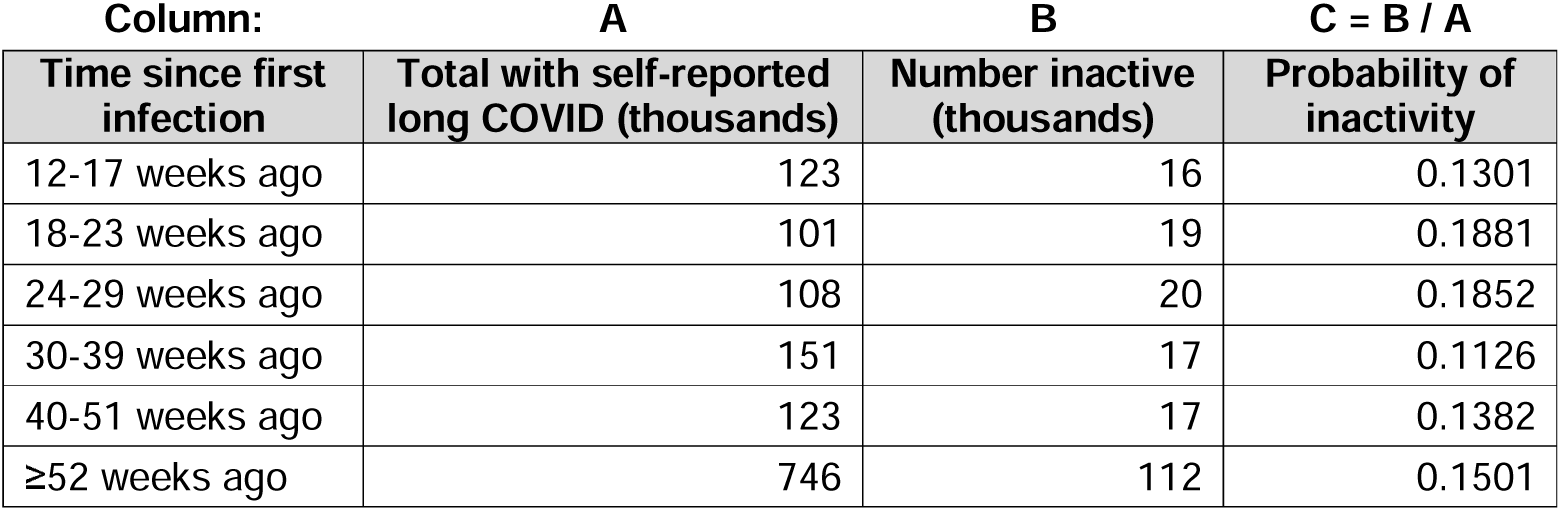
2. For each time-since-infection stratum, calculate the odds of inactivity (*o_i_*) among people reporting Long Covid from the probability: 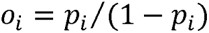 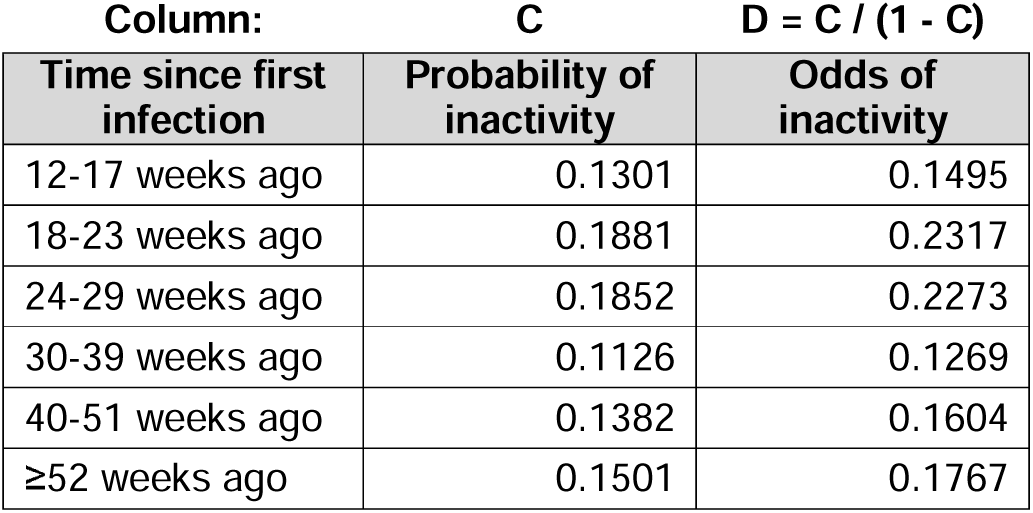
3. For each time-since-infection stratum, divide the odds of inactivity by the estimated adjusted odds ratio (aOR) for people currently reporting Long Covid (*r_i_*) in the corresponding time-since-infection stratum; this gives an estimate of the counterfactual odds of inactivity (*Õ_i_*) (that is, the odds had those reporting Long Covid not been infected with SARS-CoV-2) <u>assuming the statistical model is correct</u>: 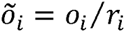 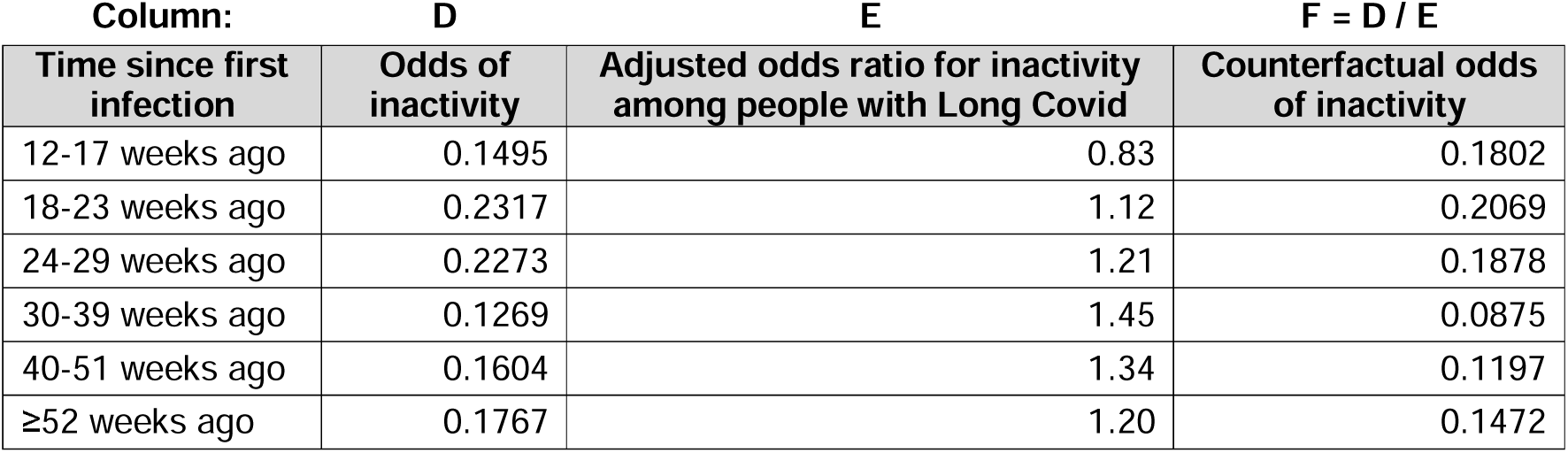
4. For each time-since-infection stratum, convert the counterfactual odds of inactivity to a counterfactual probability (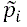): 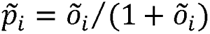 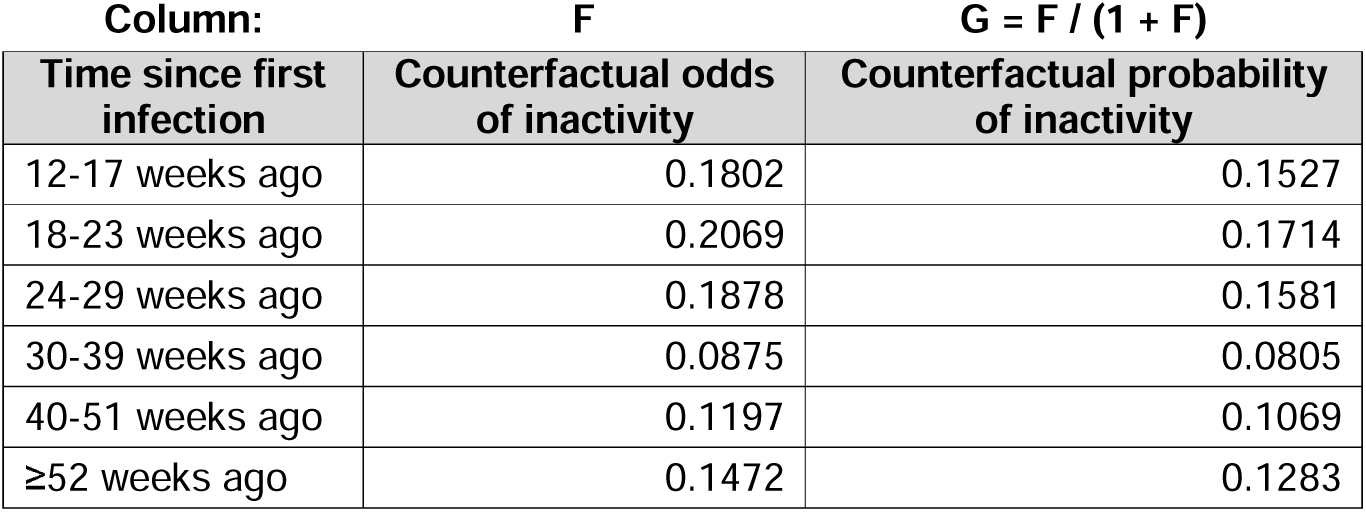
5. For each time-since-infection stratum, multiply the total number of people reporting Long Covid by the counterfactual probability of inactivity; this gives an estimate of the number of people reporting Long Covid who would have been inactive had they not been infected with SARS-CoV-2 (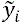) <u>assuming the statistical model is correct</u>: 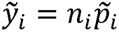 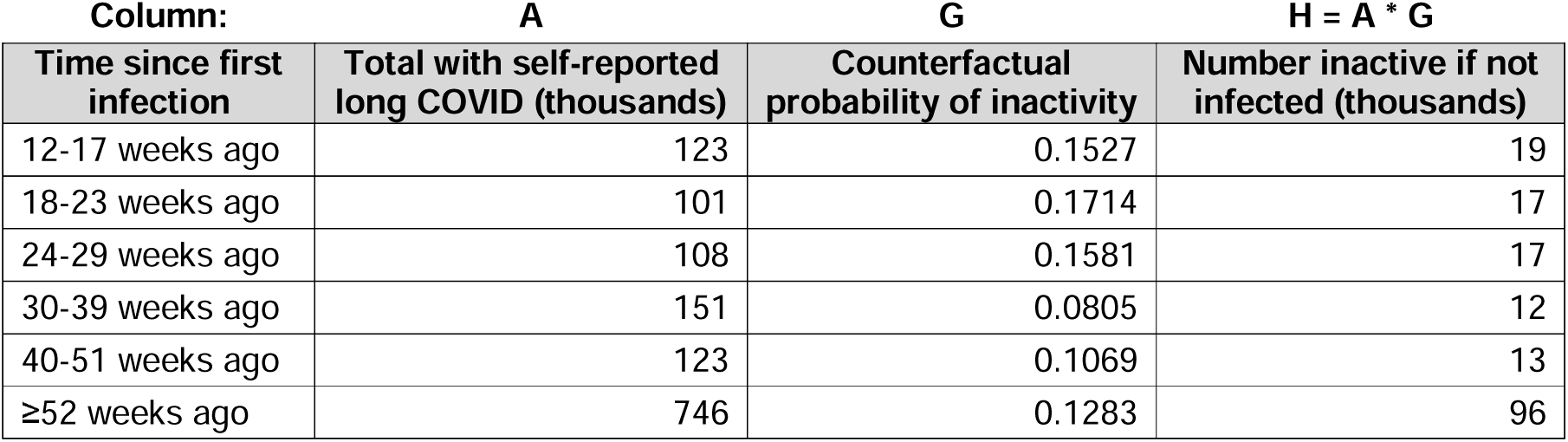
6. For each time-since-infection stratum, calculate the difference between the number of people reporting Long Covid who were inactive and the estimated number who would have been inactive had they not been infected with SARS-CoV-2; this gives an estimate of the inactivity attributable to Long Covid (Δ*_i_*) <u>assuming the statistical model is correct</u>: 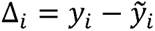 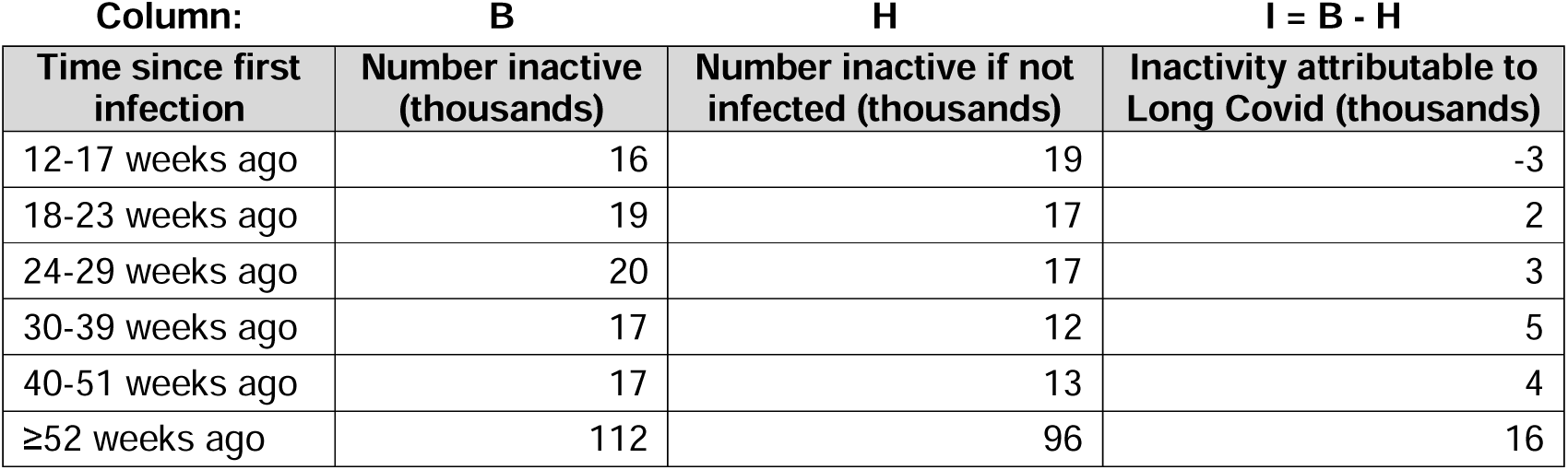
7. Sum the estimated attributable inactivity totals across time-since-infection strata: 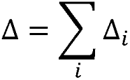 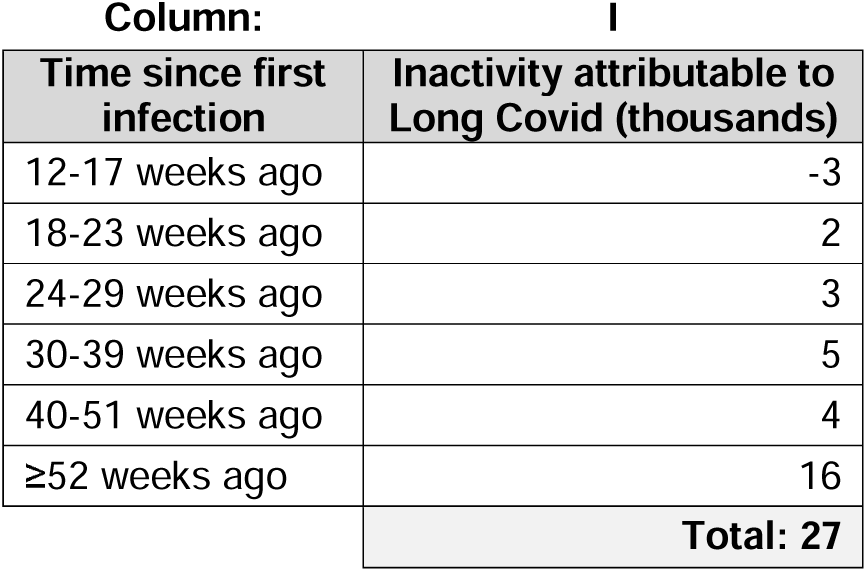

### Confidence intervals

There is uncertainty inherent in both inputs to our estimates: the number of people in the population reporting Long Covid by inactivity status; and the adjusted odds ratios for inactivity by time since first SARS-CoV-2 infection and current Long Covid status. We therefore constructed confidence intervals around our estimates using simulation:

1. For each time-since-infection stratum, take a random draw from the normal distribution with mean equal to the total number of people reporting Long Covid who are inactive, and standard deviation equal to the corresponding standard error.
2. For each time-since-infection stratum, take a random draw from the normal distribution with mean equal to the estimated coefficient for the ‘currently reporting Long Covid’ group from the conditional logit model, and standard deviation equal to the corresponding standard error.
3. For each time-since-infection stratum, take the antilog of the value randomly drawn in step 2 above to obtain the corresponding aOR.
4. Go through steps 1-7 in the ‘point estimates’ subsection, but replacing *y_i_* and *r_i_* by their randomly drawn values, 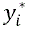 and 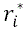, to obtain Δ*.
5. Repeat steps 1-4 above a further 9,999 times (i.e., 10,000 iterations in total).
6. Calculate the standard deviations of the sampling distributions of *y*\*and Δ*, *s^y^*\* and *s*^Δ*^ respectively. These provide estimates of the standard errors of *y* and Δ respectively.
7. Construct 95% confidence intervals around *y* and Δ:

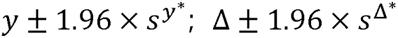

## Notes

### Author Declarations

The study received ethical approval from the South Central Berkshire B Research Ethics Committee (20/SC/0195).

### Summary of Updates

Additional explanatory text added to the Methods section; worked example of calculations added to Supplementary Appendix 2.

